# The historical fingerprint and future impact of climate change on childhood malaria in Africa

**DOI:** 10.1101/2023.07.16.23292713

**Authors:** Colin J. Carlson, Tamma A. Carleton, Romaric C. Odoulami, Cullen D. Molitor, Christopher H. Trisos

**Affiliations:** Department of Epidemiology of Microbial Diseases, Yale University School of Public Health; Public Health Modeling Unit, Yale University School of Public Health; Yale Center on Climate Change and Health, Yale University School of Public Health; Department of Agricultural and Resource Economics, University of California, Berkeley; African Climate and Development Initiative, University of Cape Town; Center for Eflective Global Action, University of California, Berkeley

## Abstract

Health-related risks from climate change are growing exponentially^1^, but direct attribution of health outcomes to human influence on the climate remains challenging ^2,3^. Here, we leverage a comprehensive dataset of 50,425 population surveys ^4^ to investigate whether human-caused climate change has increased the burden of childhood malaria across sub-Saharan Africa. In historical data, we find that prevalence shows a robust response to temperature and extreme precipitation, consistent with expectations from previous empirical and epidemiological work. Comparing historical climate reconstructions to counterfactual simulations without anthropogenic climate forcings, we find two-to-one odds that human-caused climate change has increased the overall prevalence of childhood malaria across sub-Saharan Africa since 1901. We estimate that by 2014, human-caused climate change was responsible for an average of 87 excess cases of malaria per 100,000 children ages 2 to 10 (95% confidence interval [CI]: -300, 507), with higher elevation and cooler regions in southern and east Africa experiencing greater increases. Under future climate change, we project that increasing temperatures could marginally accelerate the elimination of malaria in west and central Africa, where the present-day burden is highest, with an average overall reduction of 94 (low greenhouse gas emissions, SSP1-RCP2.6; 95% CI: -497, 160) to 1,890 (high emissions, SSP5-RCP8.5; 95% CI: -4846, 65) cases per 100,000 children in sub-Saharan Africa by the end of the century. However, we find that limiting future global warming to under 2°C (SSP1-RCP2.6) compared to 3°C (SSP2-RCP4.5) could prevent an average of 505 excess cases (95%: -199, 1209) per 100,000 children in southern Africa by 2100. Our study resolves a decades-old debate about one of the first suspected health impacts of climate change, and provides a template for future work measuring its true global burden.

## Main Text

Despite progress towards global eradication, malaria remains the single deadliest climate-sensitive infectious disease ^5^. Malaria transmission is highly responsive to temperature, driven by both the life cycle of the ectothermic mosquito vectors (*Anopheles* spp.) and the thermal sensitivity of the parasites (*Plasmodium* spp.) themselves ^6,7^. In laboratory conditions, *P. falciparum* transmission by *An. gambiae* peaks around 25°C, and becomes negligible below ∼16°C or above ∼34°C^6,7,8^. Given these biological constraints, climate change has become a major concern for populations potentially at risk of malaria in southern and high-elevation east Africa, where temperatures may no longer be prohibitive to malaria transmission ^9,10,11^. On the other hand, in west and central Africa—where the burden of malaria is highest—many studies suggest that climate change will reduce or eventually preclude transmission^9,10,12,13^.

These risks were among the first proposed health impacts of climate change ^14,15^, but have been surprisingly contentious, and even described as “hot air” ^16^ and “dangerous pseudoscience” ^17^. Malaria experts have often claimed that observed warming trends are incompatible with long-term reductions in malaria prevalence across Africa, and warned that other factors like drug resistance and funding instability pose a more serious threat to malaria eradication ^16,18,19^. Empirical evidence to test these assumptions is sparse, with the highest profile studies focusing on a single dataset of malaria incidence over several decades at a tea plantation in Kericho, Kenya. Since 2000, over a dozen studies have argued that these data either support ^20,21,22,23,24^ or undermine ^25,26,27,28,29,30,31^ the broader hypothesis that climate change is responsible for a resurgence of malaria in the east African highlands ^28,32,33,34,35^. More recently, a study by Snow *et al.*^4^ examined the last century of continent-wide changes in malaria prevalence, and concluded that observed trends could not be neatly explained by climate change, but did so based only on visual correspondence between moving averages of rainfall, minimum temperature, and modeled malaria prevalence over the entire continent.

In this study, we revisit these debates by applying state-of-the-art methods from detection and attribution, an area of climate science that quantifies the historical and real-time climate impacts of anthropogenic greenhouse gas emissions ^36,37^. These methods underpin the scientific consensus on human-caused climate change, and are regularly used to identify the role of climate change in the intensity, frequency, and distribution of specific extreme events (e.g., heatwaves, heavy precipitation, and droughts) ^38,39,40^. However, attribution remains challenging for the downstream impacts of anthropogenic climate change on people and ecosystems, and methodological frameworks for impact attribution are still comparatively underdeveloped ^3,41,42^. Applications to infectious disease dynamics are especially challenging, as relationships between climate and disease transmission are often complex, nonlinear, and confounded by human intervention, and few epidemiological datasets exist with sufficient spatial and temporal scope to resolve these relationships. As a result, hundreds of studies have tested for correlations between climate and observed changes in disease incidence or prevalence, but very few have shown that these changes are causally attributable to anthropogenic climate change ^43,2^.

Here, we draw on frameworks from climate science^38,39,40^, econometrics^44,45^, and epidemiology ^43,2^ to conduct an end-to-end impact attribution study (per ^42^), measuring the direct effect of anthropogenic climate change on long-term trends in the burden of an infectious disease. We apply this framework to estimates of *falciparum* malaria prevalence in children aged 2-10 in sub-Saharan Africa (*Pf* PR_2−10_), which experiences roughly 95% of the global burden of malaria (with 80% of deaths in children under the age of 5) ^46^. We analyze a recently published dataset with unparalleled resolution and scope (Figure 1), consisting of 50,425 surveys spanning more than a century (1900 to 2015) ^4^, which we aggregate to 9,875 monthly average values at the first administrative (state or province) level. These data capture a snapshot of population-wide prevalence at a moment in time (i.e., cases of active malaria infection per child; versus, e.g., incidence rate: new cases per child per year). Leveraging climate econometric methods^44,47,48^, we develop a panel regression model that isolates the role of temperature and extreme precipitation from other confounding factors that also shape malaria endemicity (Figure 2; see Methods for details). Nonparametric controls in the model (i.e., fixed effects) account for regional differences in seasonality, time periods with concerted elimination efforts, and other spatiotemporal variation not explained by identifiable factors, such as socioeconomic or ecological differences between populations. To quantify statistical uncertainty in prevalence-climate relationships, we repeatedly estimate the model with 1,000 spatially-blocked bootstrapped samples. We apply these models to make predictions based on 10 sets of paired historical climate simulations with and without anthropogenic climate forcing, and estimate the impact of anthropogenic climate change on malaria prevalence from 1901 to 2014 (Figure 3). Finally, we project how future climate change could further alter malaria prevalence between 2015 and 2100, based on three future climate change scenarios for low (SSP1-RCP2.6), intermediate (SSP2-RCP4.5), and high (SSP5-RCP8.5) future greenhouse gas concentrations (Figure 4).

**Figure 1:**
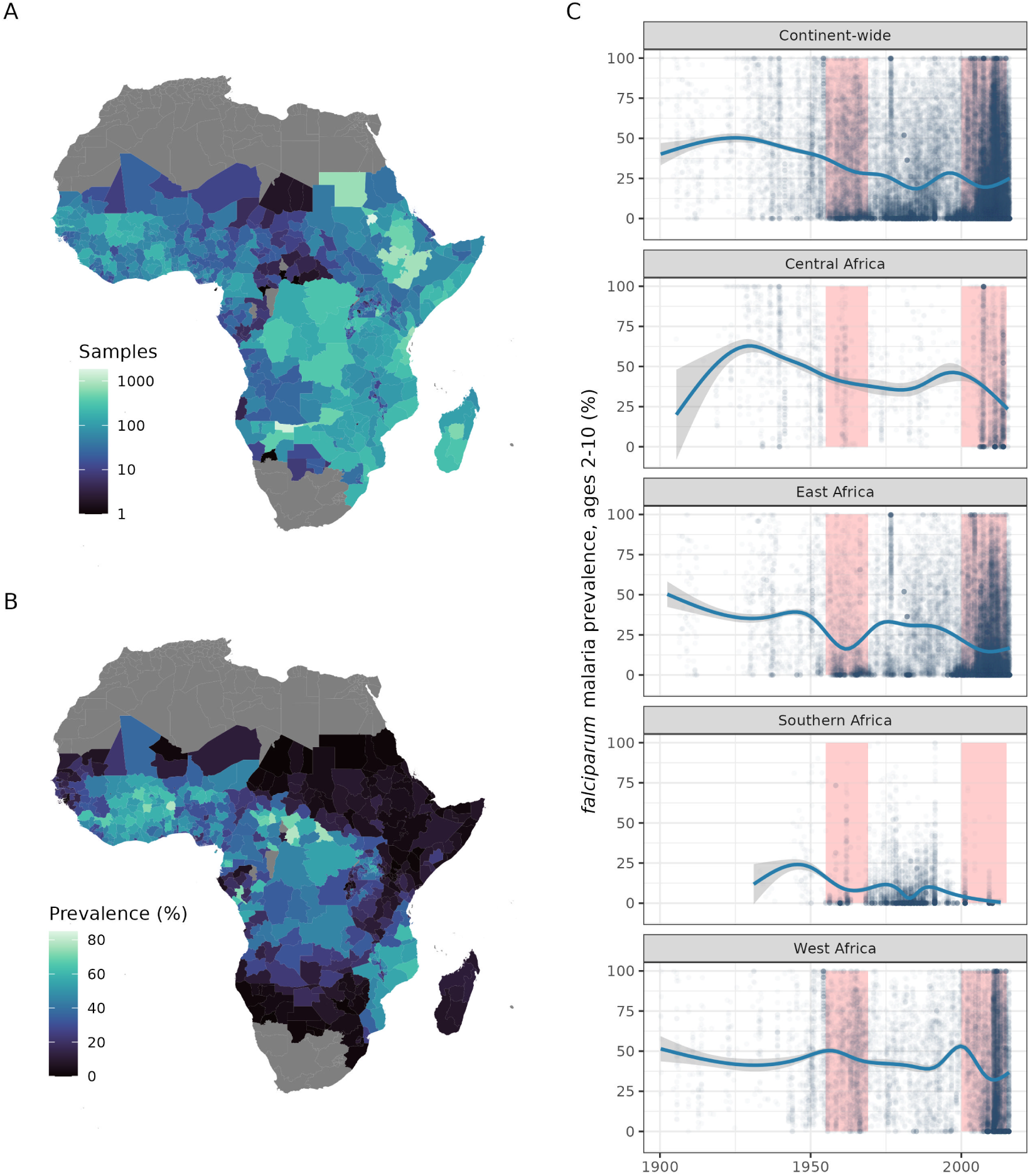
Malaria prevalence observations from 1900 to 2015. (A) The total number of malaria prevalence surveys in children ages 2 to 10 in the 20*^th^* and early 21*^st^* century, as measured by Snow *et al.* ^4^ and aggregated to the first administrative unit (ADM1). (B) Mean reported prevalence of childhood malaria over the entire sample (1900-2015, with temporal coverage varying across space). (C) Observed trends in malaria prevalence, broken down by Global Burden of Disease Study regions (see main text): each point is a single survey in the original dataset, while generalized additive models are used to construct estimated trend lines (shown in solid blue, with grey shading showing the model’s 95% confidence interval). Pink vertical bars indicate notable periods of successful malaria prevention intervention: the Global Malaria Eradication Programme (1955-1969) and the modern period including the Roll Back Malaria programs and Global Technical Strategy (2000-2014).

**Figure 2:**
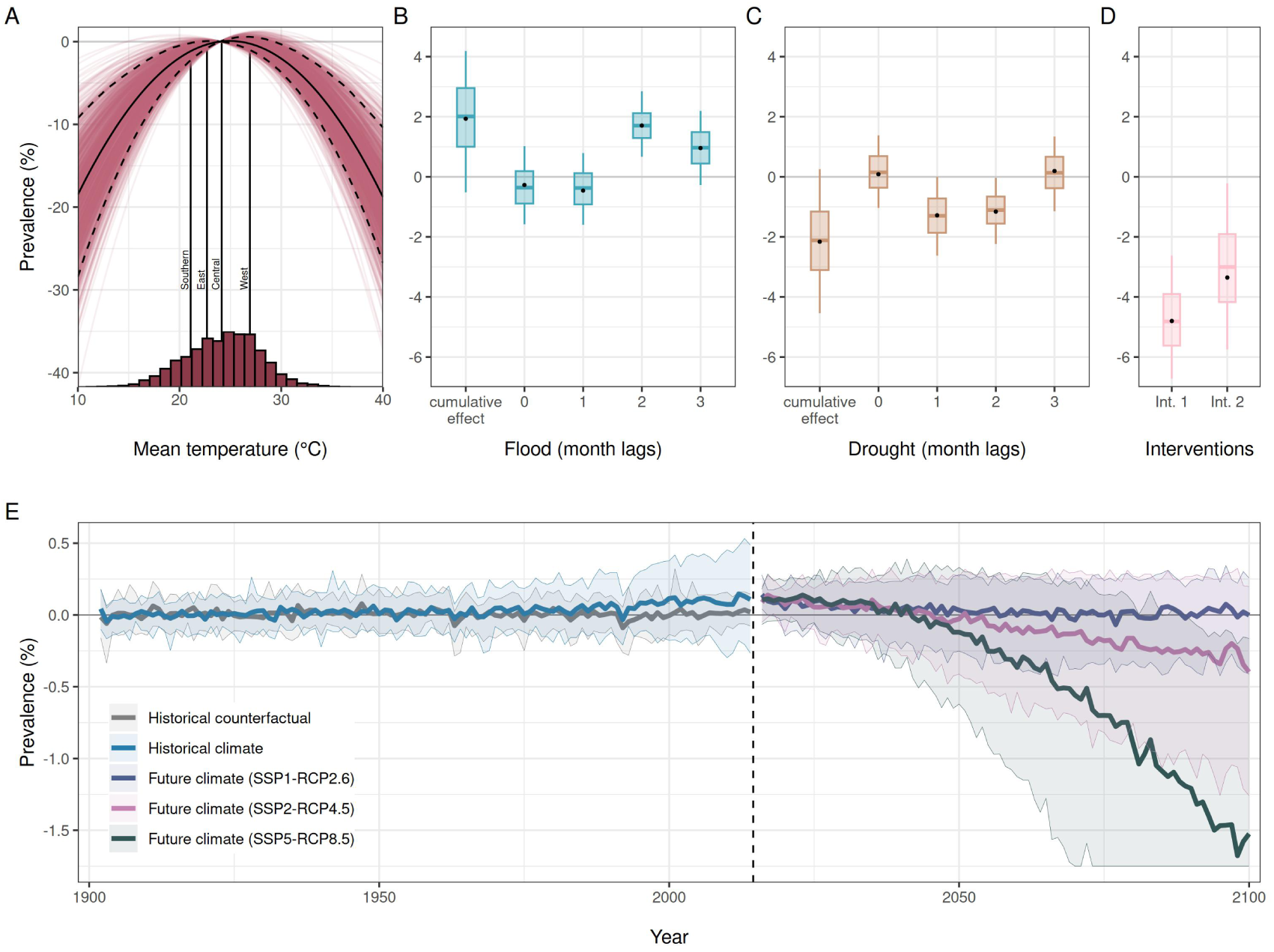
Empirical estimates of prevalence-climate relationships and predictions of climate change impacts from 1901 to 2100. (A) The estimated relationship between temperature and prevalence (point estimate in solid black; 90% CI in dashed black; bootstrapped estimates in red). Histogram shows the distribution of monthly temperatures across the estimation sample, with regional median temperatures shown as vertical lines. (B,C) The effects of extreme precipitation events (flood and drought) in the month they occur (0 lag) and after time has passed (1, 2, 3 month lags), as well as the cumulative impact across the first four months. (D) The effects of two key intervention periods (Int. 1 = 1955-1969; Int. 2 = 2000-2015) during which large-scale malaria prevention programs were implemented across the sub-continent. (B-D) Point estimates from the main model (black) are accompanied by bootstrap estimates shown as boxplots, where the box spans the 25*^th^* to 75*^th^* percentiles, the center line indicates the median, and whiskers extend to the 5*^th^* and 95*^th^* percentiles (blue, brown, pink). (E) Predicted change in prevalence attributable to anthropogenic climate change in the recent past (real historical climate given in blue; counterfactual without anthropogenic warming in grey) and in the future for low (blue: SSP1-RCP2.6), intermediate (pink: SSP2-RCP4.5), and high (green: SSP5-RCP8.5) emissions scenarios. Thick lines are the median estimates across all 10,000 simulations; shading indicates the 5*^th^* and 95*^th^* percentiles of this distribution, and is truncated at the lower axis limits for visualization purposes only (the full interval is shown in Figure S13A). Historical estimates are shown relative to an average baseline across 1901 to 1930. Future estimates are shown relative to a baseline across 2015 to 2019, added to the end-of-historical baseline (2010 to 2014). Years with incomplete predictions due to lag effects (1901 and 2015) are not displayed.

**Figure 3:**
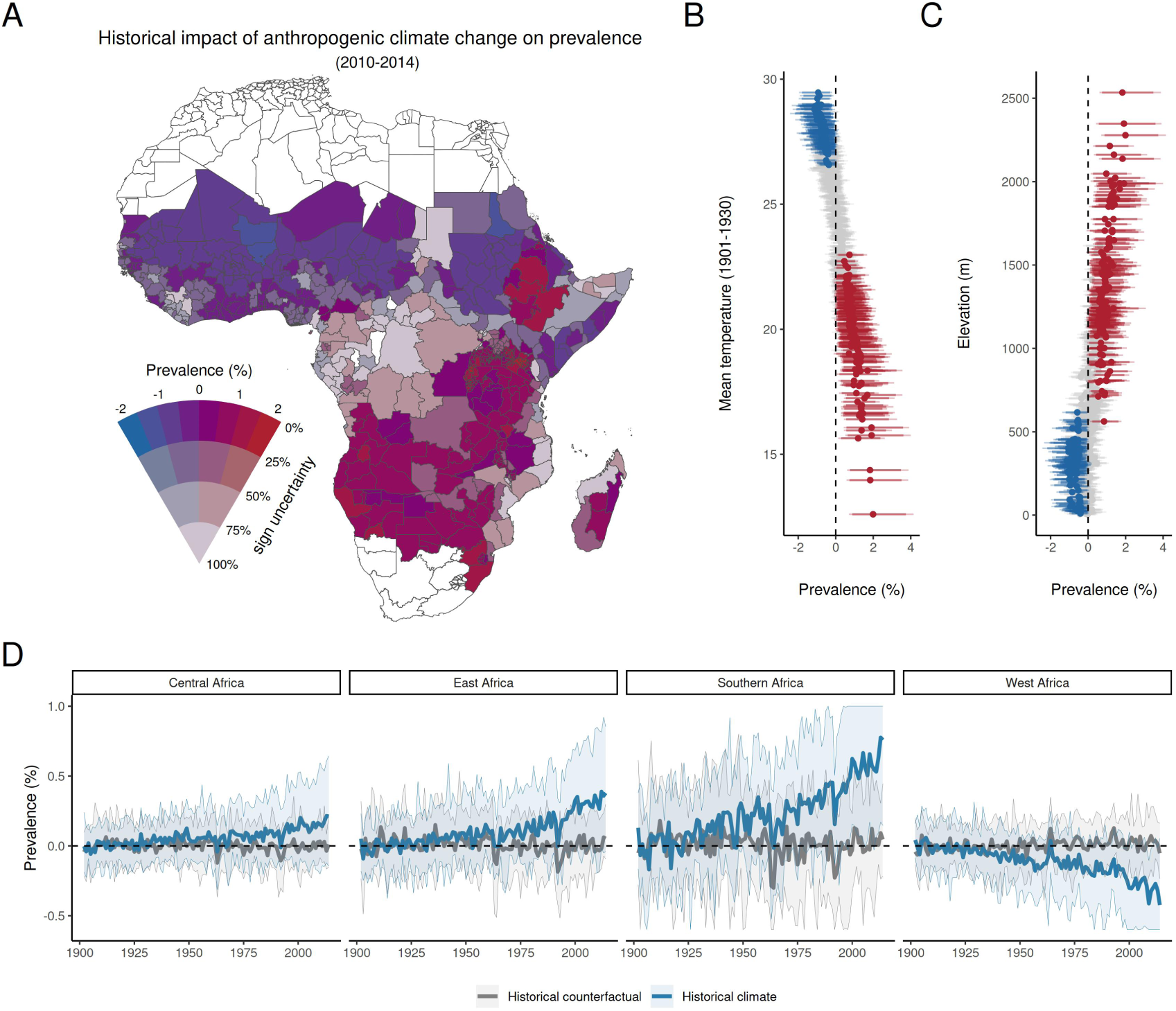
Historical changes in malaria prevalence attributable to anthropogenic climate change from 1901 to 2014. (A) Estimated change in prevalence attributable to anthropogenic climate change in each administrative unit, based on the difference between the historical climate in 2010-2014 and a counterfactual scenario for the same period simulated without anthropogenic warming. Sign uncertainty is the percentage of the 10,000 simulations that estimate an increase (for positive point estimates) or decrease (for negative point estimates) in prevalence due to anthropogenic climate change. An uncertainty of 0% implies that all models predict a positive or negative trend, while an uncertainty close to 100% indicates a near-even split of simulations showing an increase or decrease in prevalence. (B) Estimated change in prevalence attributable to anthropogenic climate change (difference between factual and counterfactual scenario in 2010-2014) in each administrative polygon, compared to the baseline mean temperature at the start of the 20*^th^* century (averaged over 1901-1930); error bars indicate both 90% (thicker lines) and 95% (thinner lines) confidence intervals. Points and lines are colored based on the 90% confidence interval: blue for negative effects, red for positive effects, and grey for non-significant effects. (C) Estimated change in prevalence attributable to anthropogenic climate change in each administrative polygon, compared to average elevation; error bars indicate both 90% and 95% confidence intervals, colored as in (B). (D) Predicted historical changes in prevalence by year, broken down by region. As in Figure 2, predictions based on true historical climate (blue) are compared to counterfactual predictions without anthropogenic warming (grey), relative to a 1901 to 1930 baseline. Thick lines are the median estimate across all 10,000 simulations; for visualization purposes, shading indicates the 90% confidence interval, and is truncated at the upper and lower axis limits. Plots begin in 1902 with the first full year of predictions (due to lag effects).

**Figure 4:**
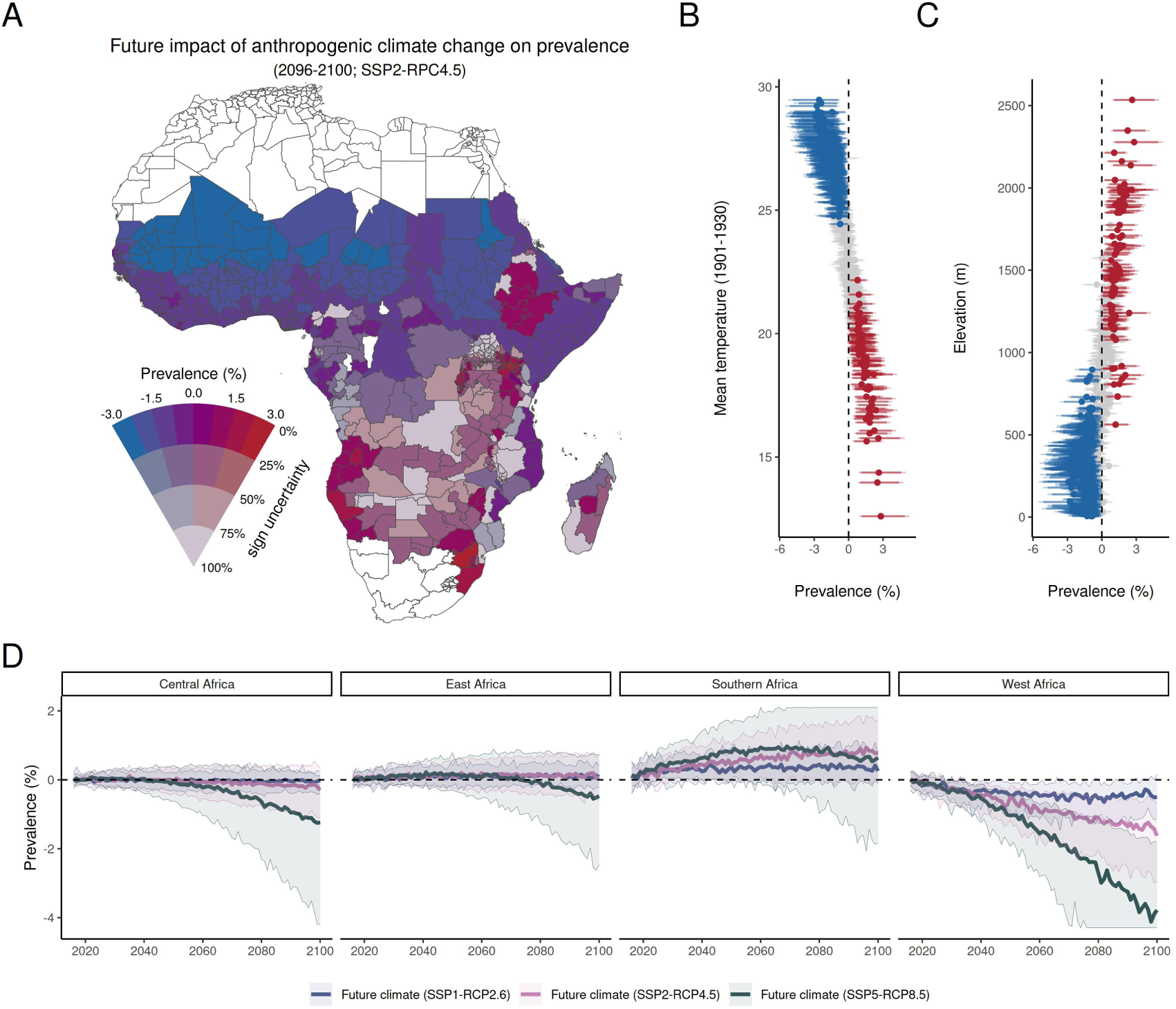
Projected future changes in malaria prevalence driven by climate change from 2015 to 2100. (A) Projected climate-driven changes in prevalence by the end of the century (2096-2100), compared to the present day (2015-2020), for an intermediate emissions scenario (SSP2-RCP4.5). Sign uncertainty is the percentage of the 10,000 simulations that estimate an increase (for positive point estimates) or decrease (for negative point estimates) in prevalence due to future climate change. An uncertainty of 0% implies that all models predict a positive or negative trend, while an uncertainty close to 100% indicates a near-even split of simulations showing an increase or decrease in prevalence. (B) Projected change in prevalence due to climate change by the end of the century (2096-2100) in each administrative polygon, estimated for SSP2-RCP4.5, compared to the baseline mean temperature at the start of the 20*^th^* century; error bars indicate both 90% (thicker lines) and 95% (thinner lines) confidence intervals. Points and lines are colored based on the 90% confidence interval: blue for negative effects, red for positive effects, and grey for non-significant effects. (C) Projected change in prevalence due to climate change by the end of the century (2096-2100) in each administrative polygon, estimated for SSP2-RCP4.5, compared to average elevation; error bars indicate both 90% and 95% confidence intervals, colored as in (B). (D) Projected changes in prevalence by year across all scenarios by region. Projections are given relative to the mean from 2015-2019, and as in Figure 2, and line color indicates scenario (blue: SSP1-RCP2.6; pink: SSP2-RCP4.5; green: SSP5-RCP8.5). Thick lines are the median estimate across all 10,000 simulations; for visualization purposes, shading indicates the 90% confidence interval, and is truncated at the upper and lower axis limits. Plots begin in 2016 with the first full year of predictions (due to lag effects).

### A robust signal of climate sensitivity

Over the last century, the prevalence of childhood malaria has exhibited a strong concave relationship with temperature (Figure 2A). Closely aligning with theoretical expectations that *P. falciparum* transmission by *Anopheles gambiae* mosquitoes should peak around 25.6°C^6^, observed values of *Pf* PR_2−10_ in our dataset peak around a monthly mean temperature of 25.8°C (Figure S1). Based on these biological expectations, we estimate the effect of temperature as a quadratic response in a panel regression model, and find that prevalence peaks at 24.9°C (95% confidence interval across the 1,000 bootstrapped models: 22.5°C, 27.0°C). These results confirm that laboratory-based studies approximate malaria epidemiology in real populations quite well, and that temperature plays a substantial role in transmission dynamics: a 10°C increase or decrease from the optimal temperature lowers prevalence by ∼8 percentage points.

The relationship between precipitation and malaria prevalence is more complex ^49^, and likely less consequential for historical trends (Figures 2B, 2C). Contemporaneous monthly precipitation exhibits a nonlinear, but highly uncertain, relationship to prevalence (see Figure S2). To parsimoniously capture nonlinear effects and disentangle divergent impacts of low and high precipitation, we define precipitation shocks with two binary indicator variables, equal to one when monthly precipitation falls below 10% (we label this a “drought shock”) or above 90% (“flood shock”) percentiles of monthly precipitation calculated for each subnational unit. While drought and flood events are complex phenomena, which develop from the combination of multiple factors (e.g., soil conditions and topography) in addition to rainfall over varying timescales, we use this drought/flood terminology as shorthand to indicate extremely low or high precipitation months. Although most effects are statistically insignificant, we find that drought shocks tend to decrease malaria prevalence 1-2 months later, while conversely, flood shocks have a positive effect on prevalence 2-3 months later. These effects and their timing are broadly consistent with expectations about how precipitation mediates the availability of mosquito breeding habitat: dry-out kills larvae and eggs, while inundation creates new breeding habitat ^50,51,52,53,54,55,56,57^. Sensitivity analyses were also weakly suggestive of another established mechanism, in which floods may wash away eggs and larvae, reducing transmission in the shorter term (see Figures S3 and S2). Overall, extreme precipitation has a measurable effect on malaria prevalence, but may be less important than temperature; however, given the sparsity of weather station data ^58^ and the uncertainty of precipitation reconstructions^59^, it is also possible that our analysis unavoidably underestimates the effect of precipitation due to measurement error. Additionally, our statistical model recovers large and highly statistically significant reductions in mean prevalence during two key intervention periods (1955-1969 and 2000-2015) that saw substantial malaria prevention programs across the sub-continent (Figure 2D).

Additional sensitivity analyses reinforce that these prevalence-climate relationships are both statistically robust and biologically consistent. Key findings are stable through time (Figure S8), and are generally insensitive to alternative model specifications, such as: the inclusion of lagged effects of temperature (Figure S4); higher-order polynomial effects of temperature (Figure S5); alternative definitions of drought and flood shocks (Figures S3, S6, and S18); controlling for the diagnostic test type (Table S1); and alternative spatiotemporal controls, which account differently for variation over space (at region, country, and state levels), time (including yearly and monthly variation), and interactions among space and time (Figure S7 and Table S2). Temperature and flood effects are strongest in rural areas (Figure S9), consistent with negative direct relationships between urbanization and malaria prevalence identified in prior work ^60,61^, as well as specific risks associated with proximity to natural water bodies or rainfed cropland in rural areas^62^. However, drought effects are highly uncertain, particularly in urban areas.

Overall, we find a robust relationship between malaria prevalence and climate, including both temperature and rainfall, that is consistent with expectations based on experimental and ecological evidence. Although climate change is unlikely to be the strongest driver of global trends in malaria endemicity (see next section), at a local scale, the month-to-month and year-to-year impacts of climate variability can be comparable in scale to the impacts of major interventions (Figure 2D). These findings suggest that malaria programs should be responsive to climate at local and national scales, and weather-based early warning systems could be useful to anticipate near-term disease dynamics ^63,64^.

### Historical impacts of climate change (1901-2014)

We find that anthropogenic climate change has, more likely than not, been responsible for a small increase in the average prevalence of childhood malaria across sub-Saharan Africa since 1901 (Figures 2E). Compared to counterfactual simulations without anthropogenic climate forcing, we estimate that by 2010-2014, anthropogenic climate change had caused an increase in continental mean *Pf* PR_2−10_ of 0.09 percentage points (*p.p.*; 95% confidence interval: -0.30 *p.p.*, 0.51 *p.p.*). Simulations with an attributable increase in continent-wide mean prevalence outnumber those with losses by two to one (proportion P_+_ of 10,000 paired factual versus counterfactual simulations with a positive difference in prevalence = 0.66). These increases are almost entirely driven by rising temperatures from anthropogenic climate forcing; the effects of drought and flood events on prevalence show no distinguishable signal from anthropogenic climate forcing over time (Figure S10). This overall trend masks substantial regional heterogeneity in historical climate change impacts (Figure 3A; Figure S11), driven almost entirely by elevational and latitudinal gradients in temperature (Figure 3B,C). For example, attributable changes in prevalence across southern Africa are high in both magnitude and certainty, with an overall increase of 0.63 *p.p.* (95% CI: -0.04 *p.p.*, 1.40 *p.p.*; P_+_ = 0.97)—nearly an order of magnitude greater than the continental mean (Figure 3D). In contrast, climate change has contributed to significantly lower malaria prevalence in west Africa (mean = -0.38 *p.p.*; 95% CI: -0.90 *p.p.*,0.02 *p.p.*; P_+_ = 0.03), where temperatures already often exceed the biological optimum for transmission. In the central African basin, a stronghold of malaria endemicity with average temperatures close to the 25^°^C optimum, the change in prevalence attributable to anthropogenic climate change is positive, relatively small, and uncertain (mean = 0.18 *p.p.*; 95% CI: -0.19 *p.p.*, 0.61 *p.p.*; P_+_ = 0.83). Finally, we estimate a meaningful overall increase in prevalence attributable to anthropogenic climate change in east Africa (mean = 0.33 *p.p.*; 95% CI: -0.14 *p.p.*, 0.87 *p.p.*; P_+_ = 0.91), but note that changes in prevalence are distributed unevenly along the steep elevational gradient: increases of up to 1-2 *p.p.* in the Ethiopian highlands and the greater Rift Valley region are accompanied by small but significant local declines throughout lowland areas in Ethiopia, Sudan, South Sudan, Eritrea, and Djibouti.

Decomposing these impacts to the monthly level reveals an interplay between space, seasons, and shifting burdens (Figure S12; Table S3). In southern Africa, rising temperatures have extended the potential tail end of the malaria season into the winter months (June and July). This supports the long-standing idea that climate change-driven poleward expansion of vector-borne diseases can emerge from shifting season lengths, and the constraints they impose on endemicity^9,10,65^. In the rest of sub-Saharan Africa, however, climate change impacts generally align with existing seasonality: for example, in lowland central and east Africa, climate change impacts are distributed much more evenly across the year, but have a stronger peak in July and August and a weaker peak in December to February ^66^. Conversely, in west Africa, the negative effects of temperature are concentrated in the hottest months (April and May), at the lowest point in the transmission cycle.

While these effects are meaningful, we caution that they are also far smaller than the reduction achieved through healthcare, mosquito nets, vector control, and economic development: previous work with the same dataset has estimated a reduction since 1900 of 16 *p.p.* (that is, a continent-wide decline in average *Pf* PR_2−10_ from 40% in 1900-1929 to 24% by 2010-2015^4^), while our estimates of historical climate change-attributable changes rarely exceed 1.5 *p.p.* for any individual administrative region. Additionally, we estimate that average reductions in prevalence realized during the Global Malaria Eradication Program (1955-1969; estimated reduction averaged over the entire period:

-4.80 *p.p.*) and recent programs like Roll Back Malaria and the Global Technical Strategy (2000–2014; estimated reduction averaged over the entire period: -3.35 *p.p.*) were substantially larger than the cumulative effects of anthropogenic climate change (Table S2). Relatively small and spatially differentiated climate-related changes in burden could have been easily concealed by the greater impact of these programs, highlighting both the success of elimination programs and the importance of using an empirical approach like ours to isolate the effect of climate from other co-evolving factors.

### Future impacts of climate change (2015-2100)

Despite contemporary trends, we project that within the next quarter-century, anthropogenic climate change will begin to reduce the prevalence of *falciparum* malaria in sub-Saharan Africa (Figure 2D; Table S4). This trend is driven largely by rising temperatures in lowland areas north of the equator, with greater possible reductions in higher greenhouse gas emissions scenarios (Figure 4). In these scenarios, temperature-related declines are slightly offset by floods, which will become more frequent across Africa ^36^, although their impact on overall trends is trivial when compared to temperature (Figure S13). Even in a future low emissions scenario (SSP1-RCP2.6: average global warming across models of +1.8°C in 2048-2052; +1.9°C in 2096-2100), increases in prevalence due to historical anthropogenic climate change are projected to essentially be offset by mid-century, stabilizing around an -0.09 *p.p.* (95% CI: -0.41 *p.p.*, 0.17 *p.p.*) projected decline across sub-Saharan Africa, relative to 2015-2020. In a high emissions scenario (SSP5–RCP8.5: +2.4°C in 2048-2052; +5.2°C in 2096-2100), we project decreases in prevalence would accelerate over time, reaching an average of -0.24 *p.p.* (95% CI: -0.80 *p.p.*, 0.26 *p.p.*) by mid-century, and -1.90 *p.p.* (95% CI: -4.85 *p.p.*, -0.07 *p.p.*) by the end of the century—a projected reduction that begins to approach the magnitude of some historical eradication programs.

Though the balance across regions will begin to shift, the geographic pattern of future changes in malaria prevalence is likely to reproduce present-day heterogeneity in impacts, as malaria transmission continues to shift along latitudinal and elevational clines in temperature (Figure 4; Figure S14). West Africa is projected to experience the most dramatic transformation, especially in a high-emissions scenario (SSP5-RCP8.5), with a projected decline of -1.04 *p.p.* (95% CI: -1.86 *p.p.*, -0.36 *p.p.*) by mid-century, and a staggering -4.25 *p.p.* (95% CI: -8.78 *p.p.*, -1.52 *p.p.*) decrease by 2100. Similar but shallower declines are projected in central Africa, where end-of-century reductions could reach between -0.06 *p.p.* (SSP1-RCP2.6; 95% CI: -0.43 *p.p.*, 0.24 *p.p.*) and -1.43 *p.p.* (SSP5-RCP8.5; 95% CI: -4.36 *p.p.*, 0.37 *p.p.*). On the other hand, localized increases in prevalence will continue in the cooler parts of the Ethiopian highlands, the greater Rift Valley region, and coastal southern Africa, potentially reaching 5 *p.p.* or more in some areas. The overall effect across east and southern Africa is a projected increase in prevalence, except in the highest emissions scenario (SSP5-RCP8.5), where both regions start to experience declines by mid-century, with east Africa eventually falling -0.60 *p.p.* (95% CI: -2.72 *p.p.*, 0.95 *p.p.*) below present-day levels by 2100.

Broadly, our results suggest that the main effect of climate change mitigation will be to keep average temperatures in sub-Saharan Africa closer to the optimum range for malaria transmission. However, for many cooler localities, such as in parts of east and southern Africa, greenhouse gas emissions reductions may prevent substantial climate change-driven increases in malaria prevalence, although uncertainty is high. For example, by mid-century, limiting global warming to below the +2°C limit in the Paris Agreement (achieved under SSP1-RCP2.6) is projected to prevent an estimated 186 cases of malaria per 100,000 children in southern Africa (95% CI: -426, 779) compared to an intermediate emissions scenario (SSP2-RCP4.5: +2.0°C in 2048-2052; +3.0°C in 2096-2100). By the end of the century, these benefits could be even greater, with 505 (95% CI: -199, 1,209) excess cases averted per 100,000 children in southern Africa (Table S4). At a more local scale, these benefits could be at least an order of magnitude greater, particularly throughout the east African highlands and parts of South Africa (Figure S14).

## Discussion

In this study, we apply an end-to-end impact attribution framework to a century of malaria surveillance, allowing us to estimate the historical and projected future impact of anthropogenic climate change on childhood malaria in sub-Saharan Africa. We find a 66% likelihood that anthropogenic climate change since 1901 has increased malaria burden; on average across Africa, an estimated 87 excess malaria cases per 100,000 people can be attributed to historical human-caused climate change. However, this burden falls disproportionately on southern and east Africa; we estimate a 97% and 91% likelihood, respectively, that anthropogenic climate change has increased present-day malaria prevalence in these regions. We project that prevalence in both southern and east Africa will remain elevated in the future: even in a low emissions scenario likely to limit global warming below +2°C (SSP1-RCP2.6), we estimate these regions will face 339 and 98 excess cases of malaria per 100,000 children by 2100, respectively. In contrast, across many other regions of Africa, we project that the overall impact of future climate change will be a net reduction in malaria: these changes are projected to be most dramatic in west and central Africa, where future climate change could reduce prevalence by up to ∼4,200 (west Africa) and ∼1,400 (central Africa) cases per 100,000 children in a high-emissions scenario (SSP5-RCP8.5). Our results suggest that climate change could be synergistic with eradication efforts in countries like Nigeria and the Democratic Republic of the Congo, where the present-day burden of malaria is highest, but will continue to create new risks in countries like Ethiopia and South Africa.

Spanning multiple centuries, our analysis is the most comprehensive look to date at the impact of climate change on any infectious disease, and brings new clarity to a decades-long debate in malaria research. Whereas some work has questioned the plausibility that overall declines in continent-wide prevalence would conceal a climate-linked increase ^4,19^, the 0.087 percentage point increase in *Pf* PR_2−10_ that we attribute to historical anthropogenic climate change could easily be masked by the nearly 200-fold greater overall reduction observed across sub-Saharan Africa over the same period. Our regional estimates also generally align with previous lab-based or site-specific empirical work, which suggests that east and southern Africa are experiencing shifts towards temperatures that are permissive to transmission for the first time or over longer seasons ^9,10^, while in west and central Africa, climate change impacts have been harder to detect, and future warming might exceed the physiological limits of malaria transmission^9,10,13,67^. Notably, our study does provide robust, empirical evidence that human-caused climate change has at least marginally contributed to malaria resurgence in high-altitude Kenya and Ethiopia, consistent with local epidemic time series or simulated dynamics based on local weather station data ^21,24,34^.

Our study therefore reconciles three long-standing ideas that are sometimes treated as paradoxical: anthropogenic climate change is not the primary force shaping past, or probably future, trends in malaria prevalence ^4,16,19^. However, anthropogenic climate change has increased the burden of malaria in sub-Saharan Africa ^14,15,21,24^, and at high elevations and latitudes, will continue to for several more decades ^9,10^. Nevertheless, rising temperatures at lower latitudes and elevations in Africa will mostly align with future efforts to eradicate *Plasmodium falciparum* from sub-Saharan Africa^10,19,67^.

In spite of climate change, elimination campaigns have already achieved substantial reductions in malaria endemicity over the last century (and not just in sub-Saharan Africa ^68^). Our study underscores that the combined benefits of disease surveillance, healthcare, vector control, and economic development can easily counter-balance climate change impacts in most places—and that malaria elimination within the next generation remains plausible, even in the face of climate change. Another recent study found that passive changes in climate, land use, and development will lead to modest reductions in malaria prevalence over the next 25 years, but with 80% effective coverage of chemotherapy, indoor residual spraying, and insecticide-treated nets, malaria could be nearly eliminated in sub-Saharan Africa by 2050^69^. In the last few years, the odds of success have become substantially higher thanks to the new RTS,S and R21 malaria vaccines: a four-dose R21 schedule could prevent between one-third and one-half of all malaria cases in children under 5^70^. Even in places where climate change is increasing malaria transmission, the combined use of classic and new interventions should have a significantly greater effect—provided that these interventions are able to continue.

At the time of writing, progress towards malaria elimination hangs in the balance, as global health financing faces an unprecedented moment of resource scarcity. Several recent anecdotes have raised relevant concerns about the fragility of elimination, such as the resurgence of malaria in Ecuador and Peru associated with migration from Venezuela ^71^, or the estimated 10,000 excess deaths due to malaria – and 3.5 million untreated cases – caused by healthcare disruptions during the 2014 Ebola virus epidemic in West Africa ^72^. Concerns about climate-linked resurgence are also more credible given the ongoing invasion of the *An. stephensi* mosquito, which thrives in cities, has already been reported in several locations in east Africa, and may be able to transmit *P. falciparum* up to much higher temperatures (∼37^◦^C) than *An. gambiae* can (∼30^◦^C)^8,73^. While our data provide suggestive evidence that temperature has historically had a *smaller* effect on prevalence in urban areas than in rural ones (Figure S9), if *An. stephensi* were to become a dominant vector across the continent, climate change might become an even more pressing concern ^74,75^. These risks only add more urgency to the global goals of eliminating both malaria and greenhouse gas emissions.

## Methods

### Malaria prevalence data

We use a recently published database of *Plasmodium falciparum* clinical prevalence in Sub-Saharan Africa^4^. This compendium, compiled by Snow *et al.* over more than two decades, is one of the most spatially and temporally complete publicly-available databases of infectious disease burden. The database covers the period from 1900 to 2015, though sampling has increased substantially since the turn of the century (pre-2000: *n* = 32, 533; post-2000: *n* = 17, 892). Most prevalence surveys used microscopy for diagnostics (*n* = 36, 805) but a substantial portion of data also derive from rapid diagnostic tests (*n* = 11, 154). The data have been compiled from a mix of archival research through public health documents, including the records of colonial governments and elimination campaigns from different periods; national survey data; electronic records published in peer-reviewed journals and grey data sources (e.g., World Health Organization technical documents); and a mix of other sources compiled by international organizations. Records were georeferenced in the original study using a standard set of protocols, with a 5km grid uncertainty threshold for point data, and broader areas stored as administrative polygons. In total, the data include a total of 50,425 prevalence surveys at a total of 36,966 unique georeferenced locations.

The Snow *et al.* data cover all available prevalence surveys, including all age ranges, but were converted by the authors of the original study to a standardized estimate of prevalence in children aged 2-10 (*Pf* PR_2−10_), using a catalytic conversion Muench model. We chose to use these standardized estimates of childhood malaria prevalence because *falciparum* malaria has the highest mortality in children and pregnant women. The trends that we infer should generally be representative of broader transmission across age groups. In some cases, we note that declines in early-life exposure can lead to smaller increases in incidence in adults ^76^; however, these impacts are likely to be small, particularly given that active and passive improvements in malaria prevention, control, and treatment much more directly determine trends in adult malaria risk.

### Climate data

We used two sets of climate data in this study. The first is an observational dataset from the Climatic Research Unit (hereafter, CRU-TS; version 4.03 for model training and bias correction), which is constructed from monthly observations from extensive networks of meteorological stations from around the globe^77^. CRU-TS provides land-only climatic variables at a high spatial resolution of 0.5° × 0.5° extending from 1901 to present (though our analysis is limited to the period 1901-2014). The second set of data is from ten global climate models (GCMs) selected from the sixth phase of the Coupled Model Intercomparison Project (CMIP6): ACCESS-CM2, ACCESS-ESM1-5, BCC-CSM2-MR, CanESM5, FGOALS-g3, GFDL-ESM4, IPSL-CM6A-LR, MIROC6, MRI-ESM2-0, and NorESM2-LM. In our historical analysis, we analysed (per GCM) one model realization of the “Historical” simulation, which includes anthropogenic greenhouse gas emissions, and one realization from the “Historical-Natural” simulation, which includes only solar and volcanic climate forcing. For both the Historical and Historical-Natural (hereafter and in the main text, “historical climate” and “historical counterfactual”) simulations, we analysed the period 1901-2014.

To investigate the continued effect of climate change on malaria prevalence between 2015 and 2100, we analysed three CMIP6 future climate change simulations from each of the 10 GCMs. Shared socio-economic pathways (SSPs) refer to the level of potential future global development (social, economic, and technological) and the implication for climate change mitigation and/or adaptation actions or policy ^78,79^. SSPs are combined with various possible future radiative forcings (representative concentration pathways; RCPs) to form the climate change scenarios used in CMIP6. Of the available SSP–RCP scenarios, we selected and used three. The first two suggest enhanced human development outcomes with increased potential towards a more sustainable (SSP1^80^) or a less sustainable (SSP5^81^) economy. The third, SSP2^82^, is a mid-way scenario, which assumes a future that mostly follows historical trends ^79^. We selected these scenarios in combination with a low (SSP1-RCP2.6), intermediate (SSP2-RCP4.5), and high (SSP5-RCP8.5) greenhouse gas concentration scenario.

We apply a standard quantile-quantile (Q-Q) bias-correction^83,84^ to the CMIP6 precipitation and temperature datasets for both of the historical simulations for the period 1901-2014, and all three future simulations for the period 2015-2100. Before the bias-correction, we first remap all simulated CMIP6 precipitation and temperature datasets to the same grid cell size (0.5° × 0.5°) as the CRU-TS observation data. We then perform for each CMIP6 model, the Q-Q bias correction at each grid-point by mapping the quantile values (*q_i_*) for the empirical cumulative distribution functions for each of the 12 months over the period 1901-2014 (for each grid point) onto the corresponding quantiles in the observational dataset (CRU-TS), so that the observed precipitation or temperature values associated with *q_i_* become the bias-corrected value in the simulations. For the counterfactual (and future) simulations, we first determine, at each grid-point, for each value of precipitation or temperature (for each month) over the period 1901-2014 (2015-2100) the equivalent quantile (*q_j_*) in the factual simulation and then identify the precipitation or temperature value associated with *q_j_* in the observational dataset as the bias-corrected value. We detrended both precipitation and temperature datasets before applying the bias-correction procedure, and then added the trends back after^84^.

### Spatial data aggregation

Our statistical analysis is designed to isolate variation in the weather that is uncorrelated with other socioeconomic and/or environmental factors that influence malaria prevalence. As detailed in the next section, we build on a large body of climate econometrics research ^45,47,85^ to do so, estimating a model that leverages variation over time in weather conditions within the same location. To estimate such a model, we require observations of malaria prevalence covering the same region in multiple time periods. In contrast, the raw prevalence data we obtain from ref. ^4^ are point data observations from individual surveys conducted at different times, such that single geolocations are not observed repeatedly over time. Therefore, we aggregate the point-level data from ref. ^4^ by averaging *Pf* PR_2−10_ observations to the first administrative level within each country (i.e., state or province level, or as shorthand, ADM1), using shapefiles provided by the Database of Global Administrative Areas dataset version 3.6 (www.gadm.org). This level of aggregation provides sufficient granularity to capture differences in climate impacts within countries and to control for local heterogeneity in confounders, while ensuring sufficient data coverage within these units. This aggregation scale was also conducted in previous work that models this dataset at the same spatial resolution ^4^. For robustness, we also show results from a statistical model that does *not* aggregate data, and instead uses the prevalence data at its native resolution (see below for details).

To compute average prevalence values at the scale of the first administrative unit (i.e., ADM1), we use an unweighted arithmetic mean over all prevalence surveys observed in the corresponding ADM1-month. This approach imposes minimal assumptions on the spatiotemporal process of malaria transmission and requires no additional high-resolution data (e.g., population) for use as weights, which are unavailable for sub-Saharan Africa for years as early as 1901. Although prior work aiming to construct comprehensive high-resolution estimates of health outcomes using point data often employs spatio-temporal smoothing methods (e.g., ref. ^86^), doing so here would artificially introduce spatial and temporal correlations that could bias recovered regression coefficients and threaten inference ^87^.

We similarly aggregate monthly 0.5^◦^ grid-level weather data(from all CRU-TS and CMIP6 models) to the ADM1-month level. To do so without introducing aggregation biases, we apply methods from previous research demonstrating that it is possible to statistically recover nonlinear relationships that take place at high spatial and temporal resolution, even when the resolution of available outcome data is relatively coarse (i.e., ADM1-month level average malaria prevalence) ^47,88,89,90,91^. In our setting, this is achieved by computing nonlinear polynomial transformations of temperature at the gridcell-by-month level *before* aggregating these values across administrative units. Such an approach ensures the temperature variables used for estimation reflect the full distribution of temperatures experienced across administrative regions of varying sizes and terrains. For example, many of Ethiopia’s 12 ADM1 regions include both hot low-elevation zones and cold highlands, such that temperatures can vary substantially within an ADM1 during the same month. We compute second-order polynomials at each grid cell before aggregating across such diverse landscapes to ensure the regressor variables capture both extreme cold and extreme heat, even when they occur simultaneously within the ADM1’s boundaries.

To see this method in practice, let *Pf PR_git_* denote average malaria prevalence in children aged 2-10 in grid cell *g* located within administrative unit *i* during month *t* and let *T_git_* indicate temperature observed at the same spatiotemporal scale. Following prior studies recovering local-level quadratic responses between malaria prevalence and temperature ^92,93^, we assume that prevalence in grid cell *g* in month *t* is a quadratic function of the temperature experienced in that same grid cell and month (noting that we show results relaxing this assumption, such as other nonlinear functional forms and the possibility of temporal lags):

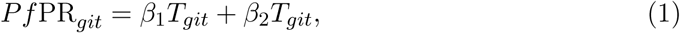

where *β*_1_ and *β*_2_ are constant average coefficients. As discussed above, we cannot empirically estimate a model like Equation 1 reliably because we do not observe prevalence over multiple months *t* for the same grid cell *g*. Instead, our empirical specification relies on average ADM1-month level prevalence variables *Pf* PR*_it_*. Thus, we must aggregate Equation 1 in a manner that allows us to recover the same *β*_1_ and *β*_2_ coefficients that describe the local-level temperature response, and which we would have recovered had we been able to estimate Equation 1 directly. Specifically, average ADM1-month prevalence can be written as:

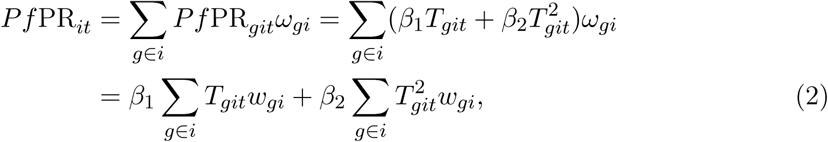

where *ω_gi_* denotes a grid-by-ADM1 weight. In our setting, we estimate an area-weighted average prevalence value by setting *ω_gi_* equal to the share of administrative unit *i*’s area that falls into grid cell *g*, as the lack of high-resolution population data make population weighting infeasible.

Equation 2 shows that a regression of average prevalence in ADM1 unit *i* and month *t* on variables that are ADM1-month weighted aggregates of the nonlinear temperature terms *T_git_* and *T_git_*^2^ will, in expectation, recover the same coefficients *β*_1_ and *β*_2_ that describe the fundamental grid-level relationship described by Equation 1. This same procedure has been used to estimate the relationship between: monthly administrative-level dengue incidence and daily grid-level temperature ^91^; annual all-cause mortality and daily grid-level temperature ^48,90^; annual country-level crop yields and daily grid-level soil moisture ^94^; among many other examples. As in these other cases, our approach mitigates aggregation bias up to the level of the grid resolution of the climate data. While climate and prevalence likely vary across space within each grid cell, we cannot resolve such dynamics here, given the lack of reliable higher-resolution weather data in Africa over the extended time frame of our analysis ^77^.

We note that one could, alternatively, construct nonlinear weather variables *after* aggregating to the administrative unit, thus estimating:

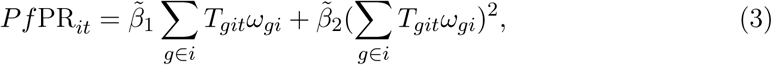

where recovered parameters *β̃*_1_ and *β̃*_2_ are biased relative to the fundamental relationship in Equation 1 because 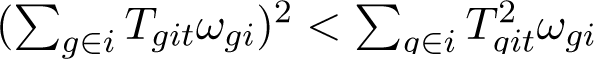, due to Jensen’s Inequality.

Thus, we construct a vector of ADM1-month temperature variables by computing nonlinear transformations at the grid level before aggregating across space, following Equation 2. While we could, in principle, follow the same procedure for precipitation, we instead compute drought and flood variables at the ADM1-month level, due to high rates of mismeasurement in grid-level rainfall estimates^77^ and due to the likelihood that prevalence-precipitation relationships occur over larger spatial scales than a single grid cell (i.e., water flows through hydrological systems linking precipitation in one location to water availability and prevalence downstream). We show in a robustness exercise detailed below that using grid-level precipitation data generates similar results as our aggregated model, but increases uncertainty, consistent with evidence on rainfall mismeasurement in Africa.

All main results rely on the spatial aggregation procedure described above. However, we additionally estimate an alternative model that leverages the point-level prevalence data directly, introducing no spatial aggregation beyond the resolution of the weather data (0.5^◦^). As we detail below and show in Supplementary Figure S15, the recovered prevalence-temperature results are very similar using these two distinct methods.

### Statistical model

The influence of climatic conditions on malaria prevalence has been heavily studied using transmission models based in vector ecophysiology and calibrated using laboratory experiments^6,7^. The important benefit of this approach is that the mechanistic links between a particular environmental condition (e.g., temperature) and malaria prevalence in the human population, such as effects on biting rate and survival probability, can be independently isolated. However, this approach is limited in its ability to generalize to real-world contexts, where complex socioeconomic factors interact with modeled relationships based on laboratory conditions. Clinical data, which measures malaria prevalence in human populations, has been used to validate modeled results^6^, but inconsistent findings arise due to challenges in statistically isolating the role of climate from the many correlated factors influencing prevalence, such as public health interventions, drug resistance, conflict and social instability, and economic shocks ^19,67,95,96,97^.

This study seeks to provide generalizable population-scale evidence of the malaria-climate link across sub-Saharan Africa using field-collected clinical data and a statistical approach designed to isolate changing environmental conditions from spatiotemporal confounding factors. Specifically, we draw on the climate econometrics literature ^47^, which has developed causal inference approaches to quantify and project the impacts of anthropogenic climate change on a host of socioeconomic outcomes, from agricultural yields^88^, to civil conflict^98^, to all-cause mortality^48^. This approach is designed to approximate controlled experiments by semi-parametrically accounting for unobservable spatial and temporal confounding factors, isolating variation in the climate system that is less likely to be correlated with other socioeconomic factors^99^. This approach is often referred to as “reduced-form”, as it allows for a plausibly causal interpretation of recovered relationships between socioeconomic conditions and the climate, but it does not easily enable the researcher to isolate individual *mechanisms* linking a changing climate to shifts in outcomes (e.g., mosquito population dynamics or parasite development rates). However, causal estimates enable counterfactual simulation in which climate is changed and all other factors are held constant; this is the exercise conducted here and in many applications of climate econometric frameworks, including estimating the impacts of climate change on dengue cases ^91^, international human migration^100^, all-cause mortality ^90,48^, and more. Moreover, these relationships can be used to calibrate more structured transmission models by providing empirical grounding from observational data.

We develop a statistical model using monthly survey-based malaria prevalence data for children aged 2 to 10 (*Pf* PR_2−10_) covering all of sub-Saharan Africa over 115 years. Our outcome variable is the average prevalence for each first administrative unit *i* (e.g., province or state) in country *c* during month-year *t*, which we denote *Pf* PR*_it_*. We estimate prevalence as a flexible function of monthly temperature and precipitation variables as follows:

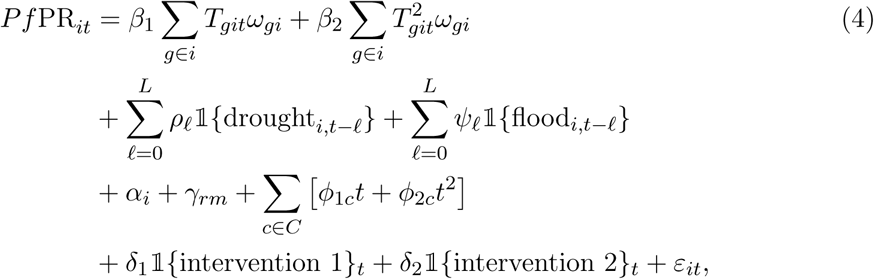

where *g* subscripts denote grid cells, which fall within administrative units *i*, and *ω_gi_* are area weights equal to the share of unit *i*’s area covered by grid cell *g*, such that Σ*_g_*_∈_*_i_ T_git_ω_gi_* equals the area-weighted average monthly temperature across all grid cells falling within administrative unit *i*. Together, parameters *β*_1_ and *β*_2_ recover a quadratic response between prevalence and monthly average temperature. As described above, polynomials are computed before aggregating across grid cells in order to preserve local nonlinearities and avoid aggregation bias. Precipitation extremes are captured by a vector of dummy variables `1`{drought*_i,t_*_−_*_l_*} and `1`{flood*_i,t_*_−_*_l_*}, which indicate whether an administrative unit’s monthly rainfall total can be categorized as drought (defined as ≤ 10% of the long-run location- and month-specific mean) or flood (defined as ≥ 90% of the long-run location- and month-specific mean) during month-year *t* − *l*. We allow for up to three months of lags (i.e., *L* = 3) for these extreme precipitation conditions in our main specification, based on hypotheses from prior literature regarding the timescales of larvae drying and of “flushing” ^50,53,101^. A variety of sensitivity analyses detailed below demonstrate that key findings are robust to: including lags for temperature as well as precipitation (Figure S4); the drought and flood cutoffs used for precipitation (Figures S3, S6, and S18); alternative functional forms of temperature (Figure S5); and the estimation of a grid-level regression that does not aggregate prevalence or weather across space (Figure S15).

Equation 4 uses a suite of semi-parametric spatiotemporal controls to isolate variation in climatological conditions that is independent from other disease transmission factors, following standard practices in the climate econometrics literature^45,47^. First, *α_i_* is a vector of indicator variables for each of 853 first administrative units (i.e., “ADM1” units) across our multi-country sample. These spatial “fixed effects” control for all time-invariant characteristics of an administrative unit that may confound the relationship between temperature, rainfall, and prevalence. For example, higher altitude regions may exhibit cooler temperatures, but they also may be composed of lower-income and more geographically isolated communities with limited access to malaria prevention interventions. By controlling for mean conditions in each location, these spatial fixed effects avoid conflating climate conditions with other geographic correlates.

Second, *γ_rm_* is a vector of region-by-month-of-year indicator variables, where regions *r* are defined using the Global Burden of Disease (GBD) regional definitions of western, southern, central, and eastern Africa (see Figure 2 in ref.^102^). Note that the subscript *m* indicates month of the year (e.g., February), while the month-year index *t* indicates the month-year time index (e.g., February, 1998). These spatiotemporal fixed effects *γ_rm_* account for region-specific seasonality in prevalence that may spuriously relate to seasonally-varying climatological conditions. We allow these seasonal controls to vary by region because of large differences in climatological seasonality and in malaria cyclicality across sub-Saharan Africa^103^, and we show below that our main findings are robust to more stringent seasonality controls defined at the country level (Figure S7). Third, *φ*_1*c*_ and *φ*_2*c*_ are coefficients estimating, for each country *c* in the full set of countries *C*, a nonlinear, country-specific quadratic in the month-year time index, which adjusts the regression for country-specific gradual trends that may confound the malaria-climate relationship, particularly under historical conditions of anthropogenic climate change. Figure S7 shows that our results are robust to multiple alternative approaches to controlling for long-run trends that may vary across space.

Finally, the indicator variables `1`{intervention 1}*_t_* and `1`{intervention 2}*_t_* are equal to one when an observation falls into the 1955-1969 or 2000-2015 period, respectively. These two periods saw substantial malaria intervention programs across the subcontinent, leading to considerable declines in malaria that were unrelated to changes in the climate ^4,104^. These indicator variables control for shocks to prevalence during these two periods, and the coefficients *δ*_1_ and *δ*_2_ allow for differential effectiveness of the two distinct intervention periods. While these variables are highly statistically significant (Table S2), our main findings are robust to their exclusion (Figure S7).

Together, these set of flexible controls imply that the residual variation in temperature and precipitation events used to identify the coefficients *β*_1_, *β*_2_, *ρ_l_*, and *ψ_l_* is month-to-month variation over time within the same location, after controlling for gradual country-specific trends, regional seasonality, and the aggregate effects of two substantial malaria prevention intervention programs.

We estimate Equation 4 using the lfe package in R. When reporting regression results directly (e.g., in Table S2), we cluster standard errors *ε_imt_* at the ADM1 level to account for serial correlation within the same region over time. When computing bootstrap samples (e.g., shown in Figure 2), we repeatedly re-estimate Equation 4 after block-resampling the full dataset using ADM1-level blocks to account for this same serial correlation. In Supplementary Table S5 and Figure S16, we show that model residuals are serially correlated, but exhibit little to no spatial correlation across ADM1 units, justifying this approach to quantifying model uncertainty (see Methods for details). However, we additionally show sensitivity to alternative methods of capturing uncertainty in Figure S17 and Table S6.

### Statistical model sensitivity and robustness

In this section, we describe a set of model sensitivity analyses that probe the robustness of our empirical model. Specifically, we investigate sensitivity of our key findings to: alternative spatiotemporal controls; inclusion of dynamic temperature effects; alternative definitions of extreme rainfall events; alternative functional forms for the prevalence-temperature relationship; and the estimation of a survey-level regression in which the point-level nature of the raw prevalence data are used directly, with minimal spatial aggregation. Finally, we provide a set of diagnostics investigating the spatiotemporal structure of our model residuals.

#### Alternative spatiotemporal controls

Our preferred empirical specification in Equation 4 includes first administrative unit fixed effects (i.e., indicator variables), region-by-month-of-year fixed effects, country-specific quadratic time trends, and two indicator variables for each of two malaria intervention periods (1955-1969 and 2000-2015). Figure S7 shows that our estimated prevalence-temperature relationship is highly robust to many alternative spatial and temporal controls. All panels in this figure include ADM1 fixed effects to control for time-invariant characteristics that may confound the relationship between prevalence and temperature, but each panel varies in the additional spatial and/or temporal controls included in the regression. A tabular version of these results is shown in Table S2. While the temperature at which prevalence peaks changes slightly across model specifications, it remains within a degree of the 24.9^◦^C value from our preferred specification for most models, particularly those including time trends that are spatially differentiated (note that peak temperatures indicated in Figure S7 are rounded to the nearest degree for display purposes). Predictably, stringent controls, such as region-by-year and country-by-month fixed effects, tend to increase statistical uncertainty. However, overall the estimated shape and magnitude of the prevalence-temperature relationship remain robust to alternative spatial and temporal controls.

#### Dynamic temperature effects

Our preferred empirical specification estimates contemporaneous (within one month) and lagged (up to three months) effects of extreme rainfall on malaria prevalence, but only contemporaneous effects of temperature. While it is possible that temperature also exhibits lagged effects, we show in Figure S4 that the cumulative effect of temperature on *Pf* PR_2−10_ is very similar whether zero, one, two, or three months of lagged temperatures are accounted for. The prevalence response to temperature does become slightly stronger with three months of lags, suggesting that our historical and future climate predictions shown throughout the main text may be somewhat conservative. However, overall these findings suggest that climate change impact predictions are unlikely to change meaningfully under different assumptions of the lag structure of temperature exposure.

#### Alternative definitions of extreme rainfall events

Our main empirical specification defines drought as months for which total precipitation is less than or equal to 10% of the long-run location- and month-specific mean. Flood is analogously defined as months for which total precipitation is greater than or equal to 90% of the long-run location- and month-specific mean. Here, we investigate the sensitivity of our main findings to these definitions. To do so, we systematically vary both the drought and flood cutoff values, ranging from *<*1% to *<*20% for drought and from *>*85% to *>*95% for flood. Figure S6 shows that the relationship between malaria prevalence and temperature is insensitive to the definition of drought and flood events. Figure S18 shows that under most drought and flood definitions, extremely low precipitation events have a negative effect on prevalence with a lag of 1-2 months. However, this effect is rarely statistically significant. Figure S3 shows that extremely high rainfall events increase prevalence with a lag of 2-3 months, a result that is statistically significant and highly robust to alternative drought and flood definitions. In general, these sensitivity analyses show that our main findings are not sensitive to the specific definitions of drought and flood used in estimation of Equation 4.

#### Alternative functional forms for the prevalence-temperature relationship

Following from theoretical and laboratory-based literature (e.g., refs.^6,7^), we model the prevalence-temperature relationship as quadratic. However, Figure S5 shows that this relationship is similar when more flexible functional forms are used. In particular, the temperature at which prevalence peaks changes little when higher order polynomials are estimated. Estimating higher order polynomials increases uncertainty, particularly in the tails of the temperature distribution, but point estimates are similar across the majority of the observed temperature range.

#### Estimating a grid-level regression

As described above, our main analysis relies on an ADM1-level regression in which malaria prevalence and weather data are aggregated from higher spatial resolutions to the ADM1 scale. Here, we show the results from an alternative approach, in which point-level malaria prevalence survey observations are matched to corresponding 0.5^◦^ resolution CRU climate data grid cells and the regression is estimated at this grid level.

Using these disaggregated data, we estimate a regression model analogous to Equation 4, but modified to fit the spatial scale of the data. Specifically, we estimate:

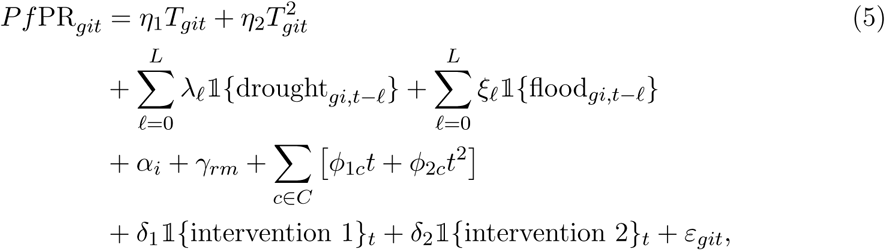

where all variables are defined as above for Equation 4. In particular, *Pf* PR*_git_* represents prevalence reported for a survey located in grid *g* falling within ADM1 unit *i* during month *t*, *T_git_* denotes temperature in the same grid and month, and precipitation extremes are captured via dummy variables `1`{*drought_gi,tl_*} and `1`{*flood_gi,tl_*} that indicate when each grid cell’s monthly rainfall total is less than 10% of its long-run month-specific mean (drought) or more than 90% (flood).

Two features render this estimating equation distinct from the main analysis. First, temperature and precipitation variables are matched exactly to the grid cell within which the malaria survey was conducted, such that no aggregation is necessary. This increases the precision of the match between weather and outcome variables. Second, the spatial “fixed effects” denoted by *α_i_* – that is, indicator variables for each of the ADM1 units in our sample – are estimated at a lower spatial resolution (ADM1) than the data itself (grid). This implies that the weather variation used to identify coefficient vectors ***η***, ***λ*** and ***ξ*** includes both variation over time within an ADM1, but also across survey locations located within the same ADM1. Thus, while this model has the benefit of leveraging high spatial resolution in weather, it potentially suffers from omitted variables bias, as weather conditions in different survey locations may be correlated with other unobservable determinants of malaria prevalence (e.g., access to health care, rates of poverty, proximity to water bodies).

We show in Figure S15 that the malaria prevalence-temperature relationship recovered from estimation of Equation 5 is incredibly similar to that estimated from our main regression model in Equation 4, suggesting that aggregation of the underlying survey data does not influence the key results of the paper. In contrast, the estimated drought and flood coefficients recovered from the grid-level regression are highly imprecise, consistent with substantial measurement error in local-level precipitation datasets in Africa for much of the 20*^th^* century ^77^.

#### Spatiotemporal structure in model residuals

We evaluate the extent to which our model residuals are correlated over space and time using two tests. First, we calculate correlations between residuals across various subsets of our data, following similar tests in ref. ^105^. Specifically, we calculate correlations across observations that are: (i) within the same ADM1 unit but from different time periods; and (ii) from different ADM1 units but within the same time period. For temporal correlations, we compute correlations between temporally consecutive observations within up to 3 months of lags, while for spatial correlations we investigate correlations within countries, across countries, within Global Burden of Disease multi-country regions, and based on physical distance (using ADM1 centroids).

Table S5 reports mean correlations, as well as the first and third quartiles of the distribution of correlations across different pairs of units. Rows labeled “temporal” quantify correlations over time while rows labeled “spatial” quantify correlations over space. We note that these tests should be interpreted with care, as the malaria prevalence data are highly unbalanced in space and time, often leaving few observations with which to estimate correlations and/or few regional or temporal pairs over which to summarize a distribution of correlations. To ensure interpretability, we restrict analysis to correlations with at least 10 observations. The last column in Table S5, labeled *N*, indicates the number of pairs for which sufficient data were available to construct correlations for a given grouping. These results reveal moderate serial correlation in residuals, especially for consecutive observations (row 2) (mean *ρ* = 0.40 for consecutive month-years within the same ADM1 unit). In contrast, spatial correlations range from negligible (mean *ρ* = 0.04 across ADM1s from different countries or regions) to modest (mean *ρ* = 0.15 and *ρ* = 0.30 for ADM1s with centroids greater than 500 km and less than 500 km of one another, respectively).

Second, we use a higher-powered test to flexibly investigate spatial correlation by estimating an empirical variogram of our residuals. A variogram estimates the variance of residuals within groups of observations as a function of the distance between them. This is a standard spatial statistics method to systematically assess spatial correlations across a sample ^106^. Figure S16 shows the resulting variogram for raw values of the outcome variable (a) and model residuals (b). Each panel shows semi-variances on the *y*-axis and distance between centroids of ADM1 units on the *x*-axis. When spatial correlation is present, semi-variances rise with distance, as in panel (a). In contrast, panel (b) shows essentially a flat line, indicating little to no spatial correlation in model residuals.

Given these results, we cluster standard errors at the ADM1 level, accounting for correlation in model residuals across all month-years for a given administrative unit. However, in Figure S17 and Table S6, we show sensitivity of our recovered confidence intervals to alternative approaches to standard error estimation, including those that account for spatial correlation (e.g., Conley standard errors).

### Predictions

In both historical and future simulations, we apply the estimated panel regression to calculate the effect of climate change on *Pf* PR_2−10_. Our predictions capture the full range of statistical uncertainty (1,000 bootstrapped model estimates) and climate model uncertainty (10 climate models), producing a total of 10,000 estimates of historical or future impacts in any given scenario. Each of these 10,000 estimates is normalized to a long-run baseline (past: 1901-1930; present: 2015-2020) before estimates are averaged, creating an estimate of climate change impacts relative to that baseline. In our historical analyses, we only use these models to estimate changes in prevalence attributable to climate change: while the panel regression model accounts for other historical drivers through the fixed effects structure, these are not the focus of our analysis, and so we choose not to estimate total prevalence including these effects. Similarly, we elect not to make assumptions about non-climate drivers of malaria prevalence in the future, and thus do not apply the model to predict future trends in overall prevalence.

For overall trends (e.g., reported in Figures 2D, 3D, and 4D), we generated continent-wide averages or four regional averages using the unweighted average of estimates for each ADM1 unit. This is a deliberate oversimplification, as we do not adjust averages based on either ADM1 units’ land area or the estimated population they contain; we made this decision based on the challenges of reconstructing historical population density at fine scales, as well as the need to otherwise make assumptions about how disease burden is allocated over space (e.g., the distribution of transmission across rural or urban areas). For similar reasons, we chose not to estimate the effect of prevalence changes on overall malaria incidence. Although some studies have attempted this using a linear conversion with total population^107^, proper estimation of incidence (and the effects of treatment variables, through prevalence, on case burden) requires malaria transmission models that require substantially more demographic assumptions ^104^. Future work could explore both of these methodologically-complex directions, and potentially generate finer-scale estimates of how many cases of childhood malaria, and resulting deaths, are attributable to climate change.

## Data Availability

All data used in the study are available from other sources. All code are available on Github.

http://www.github.com/cjcarlson/falciparum

## Acknowledgements

We thank Sadie Ryan and Rory Gibb for thoughtful conversations that supported this work, and Jonathan Proctor for constructive feedback on the manuscript. CJC was supported by a Yale School of Public Health Transformation Award. CHT was supported by the University of Cape Town Future Leaders Programme and by the FLAIR Fellowship Programme: a partnership between the African Academy of Sciences and the Royal Society funded by the UK Government’s Global Challenges Research Fund. RCO was supported by the Carnegie Corporation of New York through the Development of Emerging Academic Leaders (DEAL) in Africa and the German Academic Exchange Service (DAAD) ClimapAfrica programme.

## Data Availability

No original data are generated or reported in our study. All data used in analyses, including both malaria and climate data, is freely available and referenced in the Methods. For replication purposes, we make our pre-processed dataset available at TBD. Climate model simulation data are too large for standard public data repositories, but are available upon request.

## Code Availability

All code is available at github.com/cjcarlson/falciparum.

## Supplementary Results

### Supplementary Figures

**Figure S1:**
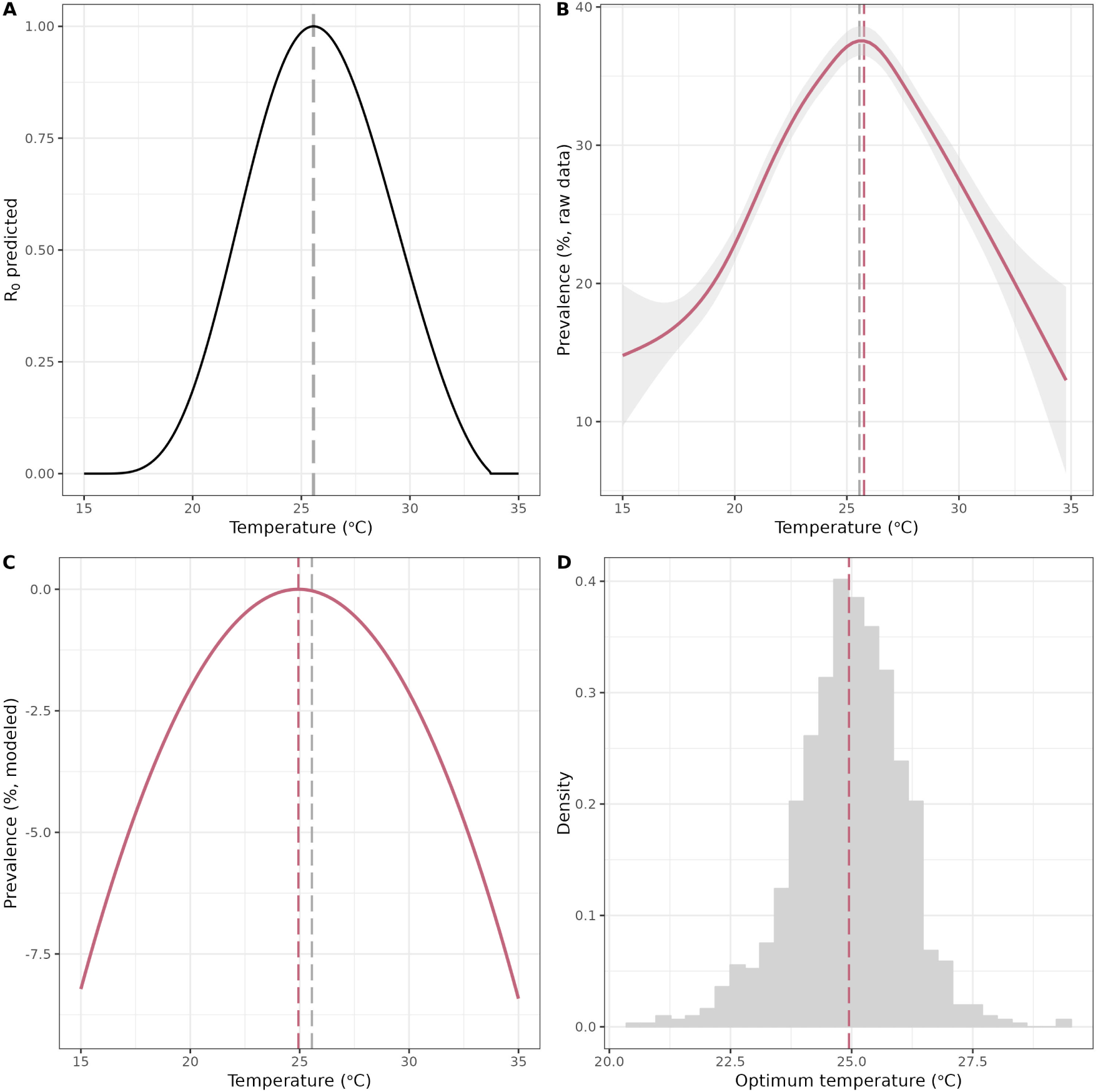
Malaria prevalence follows biological expectations. (A) The theoretical expectation for *R*_0_(*T*), the scaled partial response of the basic reproduction number to temperature, estimated based on laboratory experiments (black line) ^6^. Transmission peaks around an estimated optimum of 25.6 *^◦^*C (grey dashed line). (B) Observed malaria prevalence data from Snow *et al.* ^4^ matched to monthly temperature from CRU weather station data, summarized and smoothed using a generalized additive model. The observed optimum temperature (red dashed line) closely matches expectations based on laboratory experiments (grey dashed line). (C) Main panel regression estimate for prevalence response to temperature (also shown in Figure 2A). The modeled optimum temperature (red dashed line) is slightly lower than in laboratory experiments (grey dashed line). (D) Histogram of optimum temperatures derived from 1,000 bootstrapped estimates of the panel regression model shown in panel (C). The mean optimum temperature across all bootstrap samples (red dashed line) is identical to the optimum shown in panel C.

**Figure S2:**
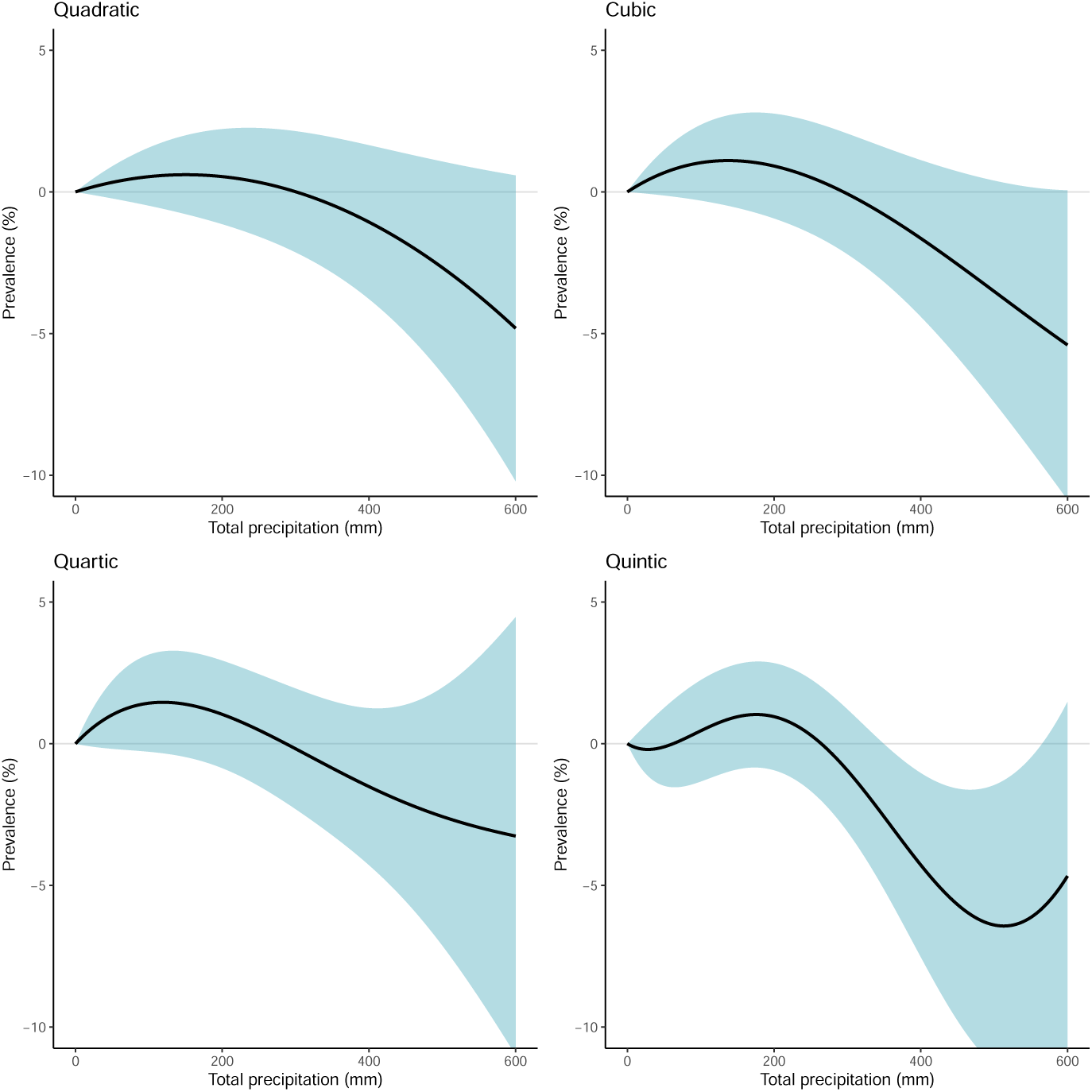
Non-linearities in the *Pf* PR_2_*_−_*_10_-precipitation relationship. All panels show the estimated relationship between malaria prevalence for children aged 2-10 and monthly cumulative precipitation and are plotted relative to a month with no rainfall. All relationships are estimated for contemporaneous monthly precipitation. All standard errors are clustered at the first administrative unit (e.g., province) level.

**Figure S3:**
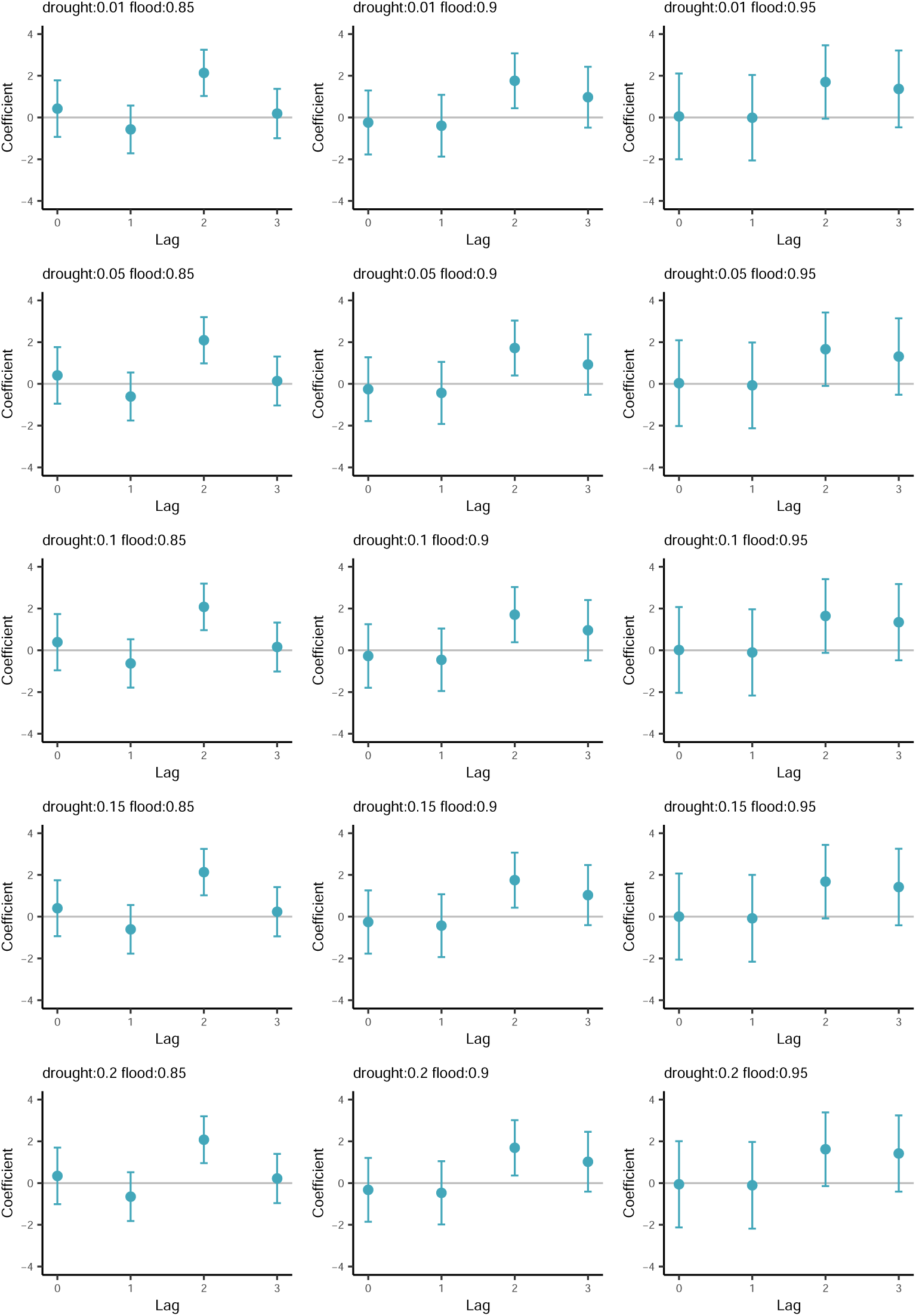
Sensitivity of *Pf* PR_2_*_−_*_10_-flood relationship to alternative drought and flood definitions. All panels show the estimated relationship between malaria prevalence for children aged 2-10 and contemporaneous and lagged flood events. Point estimates are given by solid circles, while vertical bars indicate 95% confidence intervals. Cutoff values for drought and flood definitions are given in the titles of each panel. For example, the first panel in the upper left defines drought as monthly total precipitation that falls below 1% of the long-run location- and month-specific mean, and defines flood as monthly total precipitation that falls above 85% of the long-run location- and month-specific mean. In all specifications, three months of lagged precipitation extremes are included, as well as all other controls shown in Equation 4. The main specification used throughout the paper uses a drought cutoff of 10% and a flood cutoff of 90%. All standard errors are clustered at the first administrative unit (e.g., province) level.

**Figure S4:**
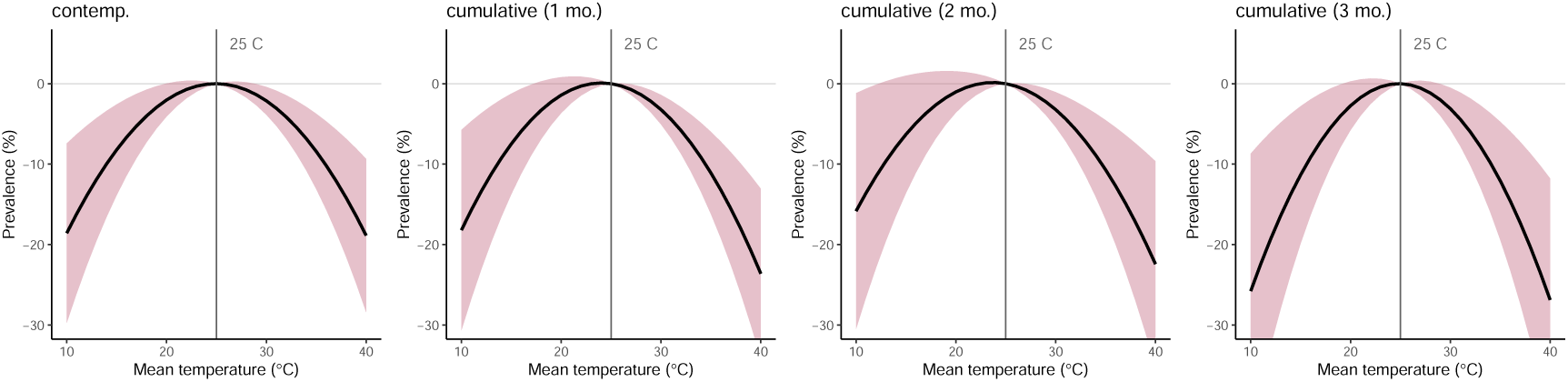
Cumulative effect of contemporaneous and lagged temperature on *Pf* PR_2_*_−_*_10_. All panels show the estimated relationship between malaria prevalence for children aged 2-10 and monthly average temperature and are plotted relative to a monthly average temperature of 25*^◦^*C. The first panel shows the effect of monthly average temperature on the same month’s average prevalence (this is the main estimate used throughout the main text). The second panel shows the cumulative effect of contemporaneous temperature and the prior month’s temperature on prevalence, while the last two columns show analogous results for two and three months of lags, respectively. In all specifications, three months of lagged precipitation extremes are included, as well as all other controls shown in Equation 4. All standard errors are clustered at the first administrative unit (e.g., province) level.

**Figure S5:**
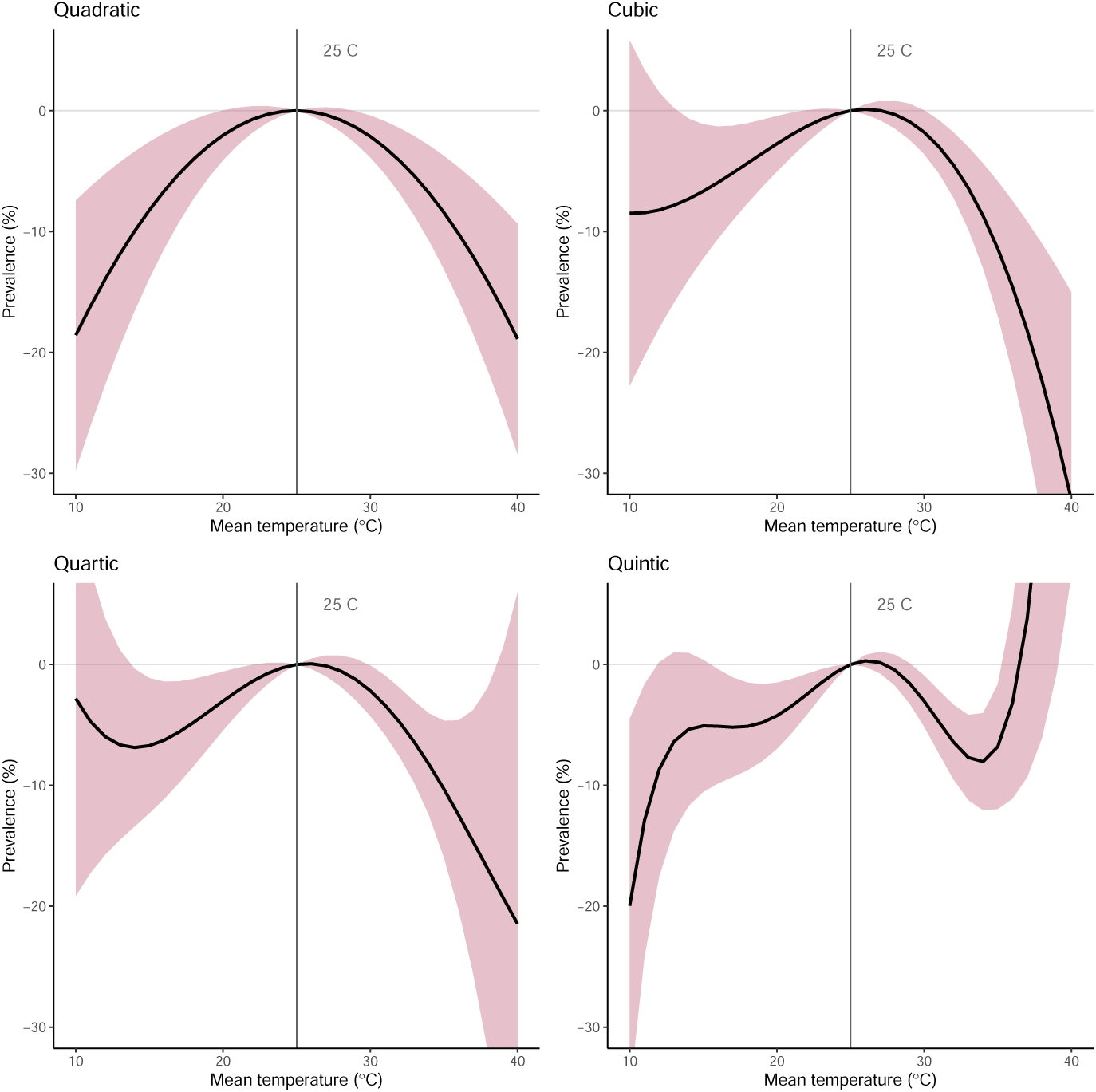
Alternative functional forms for the *Pf* PR_2_*_−_*_10_-temperature relationship. All panels show the estimated relationship between malaria prevalence for children aged 2-10 and monthly average temperature and are plotted relative to a monthly average temperature of 25*^◦^*C. Starting in the upper left, the first panel shows the paper’s main specification, a quadratic functional form for the prevalence-temperature relationship. The second panel shows a cubic functional form, the third a quartic, and the fourth a quintic. All standard errors are clustered at the first administrative unit (e.g., province) level.

**Figure S6:**
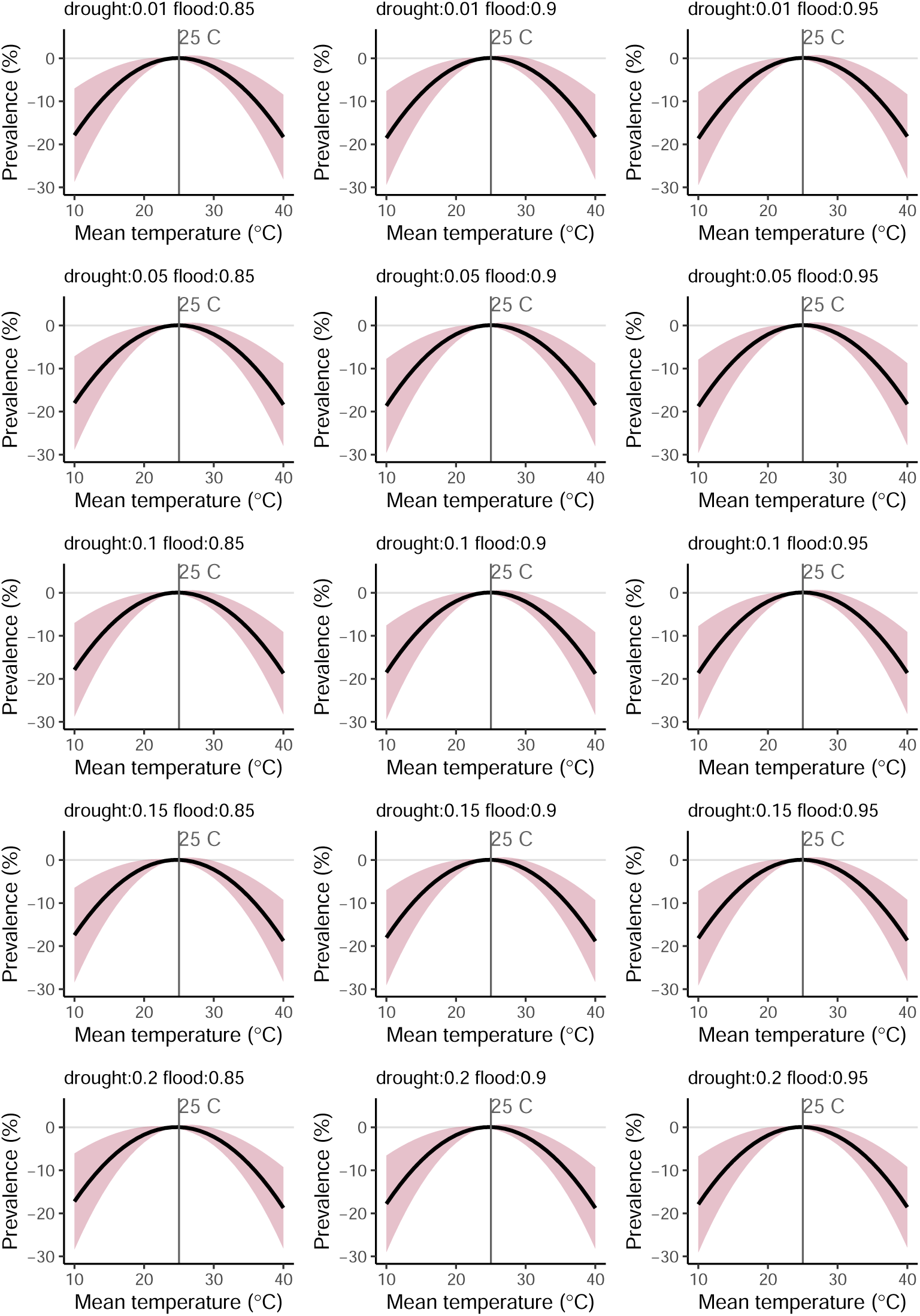
Sensitivity of *Pf* PR_2_*_−_*_10_-temperature relationship to alternative drought and flood definitions. All panels show the estimated relationship between malaria prevalence for children aged 2-10 and monthly average temperature and are plotted relative to a monthly average temperature of 25*^◦^*C. Cutoff values for drought and flood definitions are given in the titles of each panel. For example, the first panel in the upper left defines drought as monthly total precipitation that falls below 1% of the long-run location- and month-specific mean, and defines flood as monthly total precipitation that falls above 85% of the long-run location- and month-specific mean. In all specifications, three months of lagged precipitation extremes are included, as well as all other controls shown in Equation 4. The main specification used throughout the paper uses a drought cutoff of 10% and a flood cutoff of 90%. All standard errors are clustered at the first administrative unit (e.g., province) level.

**Figure S7:**
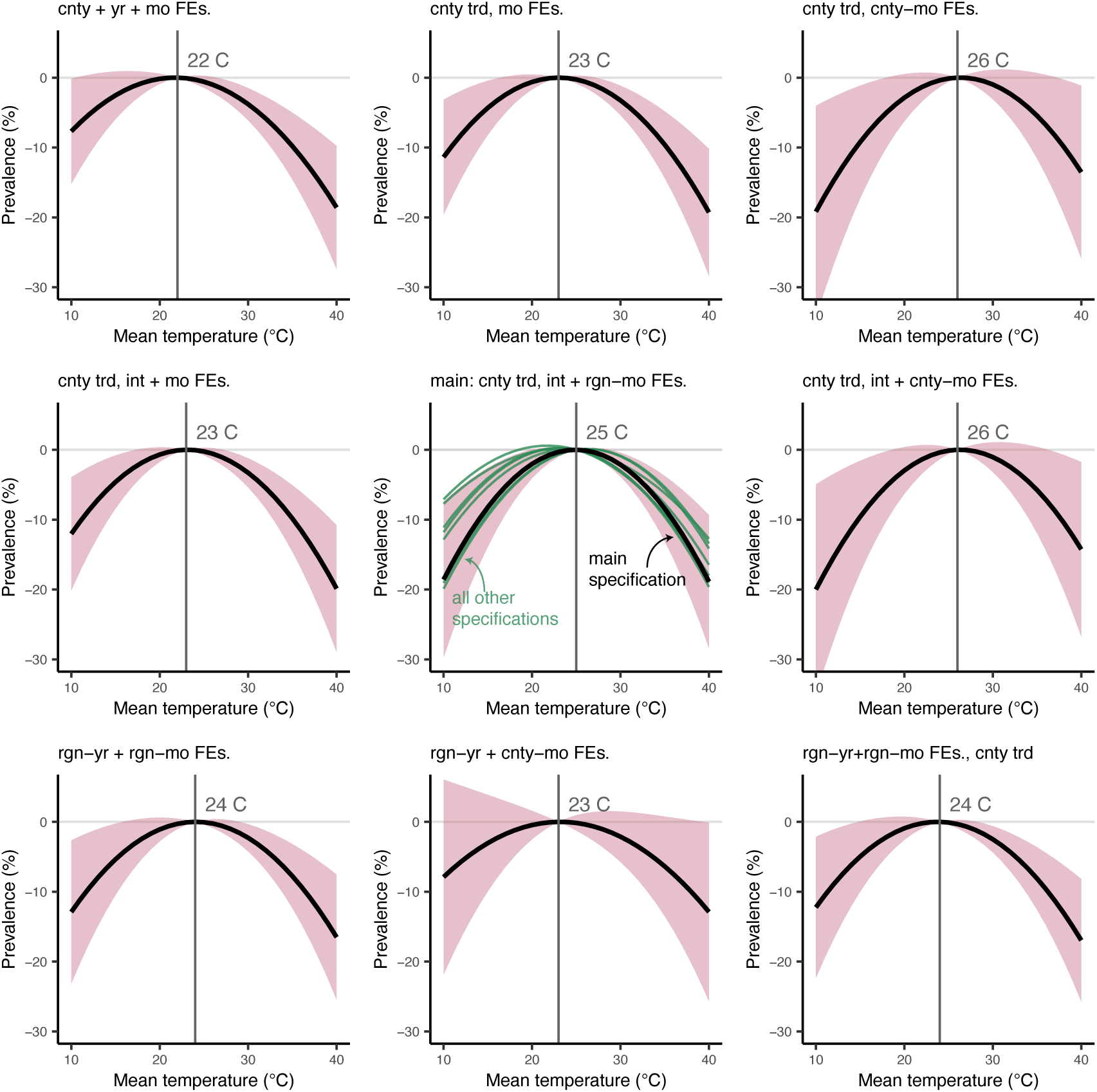
Sensitivity of the *Pf* PR_2_*_−_*_10_-temperature relationship to alternative spatiotemporal controls. All panels show the estimated relationship between malaria prevalence for children aged 2-10 and monthly average temperature and all include fixed effects (i.e., dummy variables) at the scale of the first administrative unit (i.e., ADM1). All temperature responses are plotted relative to the model-specific temperature at which prevalence is maximized; this peak temperature is indicated in grey text and with a vertical grey line in each panel. From top-left to bottom-right, model controls are: country, year, and month fixed effects; country-specific quadratic time trends and month fixed effects; country-specific quadratic time trends and country-by-month fixed effects; country-specific quadratic time trends and intervention period and month fixed effects; country-specific quadratic time trends and intervention period and region-by-month fixed effects; country-specific quadratic time trends and intervention period and country-by-month fixed effects; region-by-year and region-by-month fixed effects; region-by-year and country-by-month fixed effects; and region-by-year and region-by-month fixed effects and country-specific linear time trends. The preferred specification used throughout the main text is the center panel, which includes country-specific quadratic time trends and intervention period and region-by-month fixed effects. In this center panel, the main specification is shown in black (with pink shaded 95% confidence intervals) and all alternative specifications from other panels are overlaid in green. In all panels, “region” refers to the Global Burden of Disease regional definitions of western, southern, central, and eastern Africa. All standard errors are clustered at the first administrative unit (e.g., province) level.

**Figure S8:**
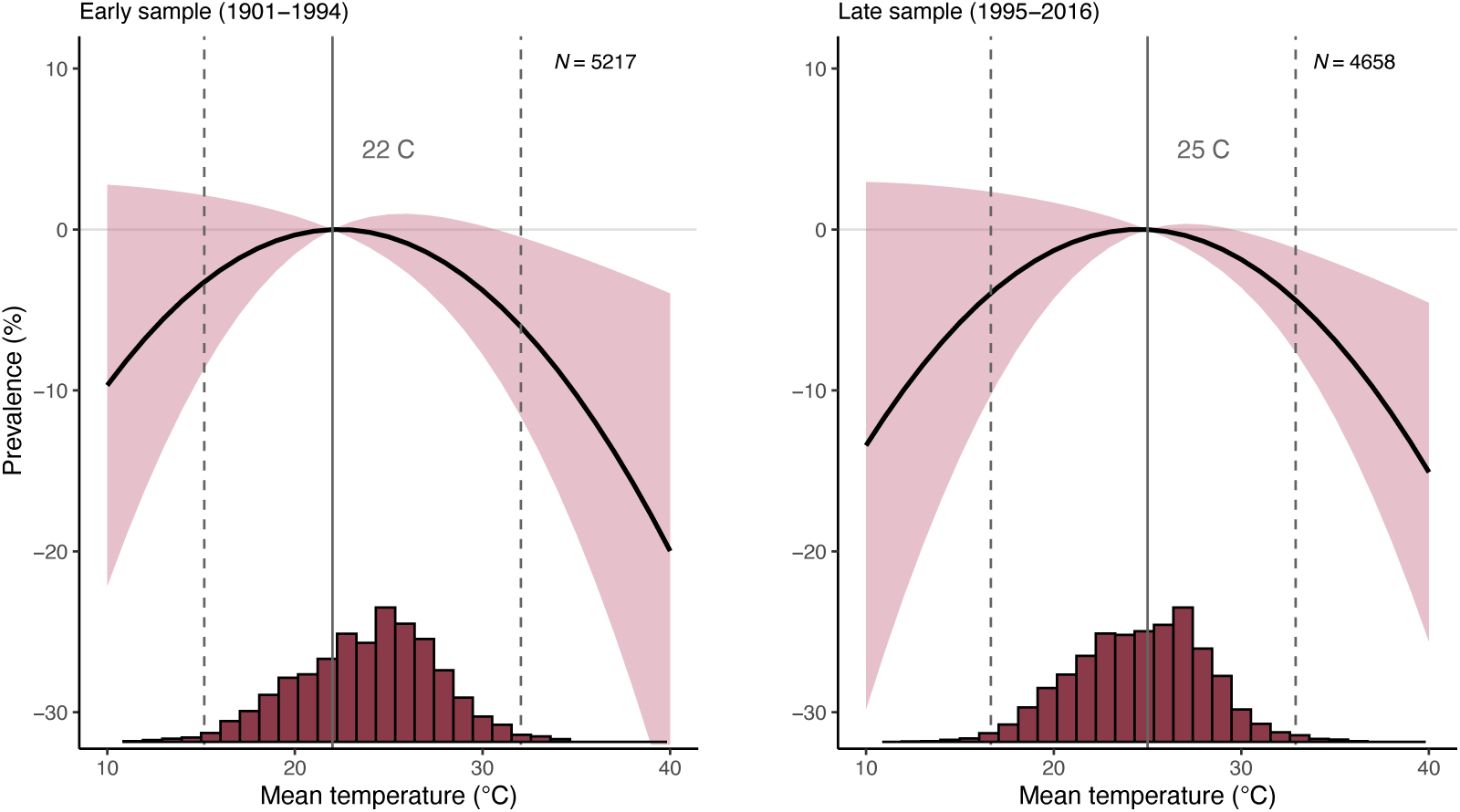
Heterogeneous effects of temperature on malaria prevalence over time. Panels show the estimated relationship between malaria prevalence for children aged 2-10 and monthly average temperature along with a histogram of the associated mean monthly temperature data with the 1st and 99th percentiles of the temperature distributions shown as dashed vertical lines. (Left) The relationship between prevalence and mean monthly temperature for years 1901 to 1995. (Right) The relationship between prevalence and mean monthly temperature for years 1995 to 2014. Both regressions include fixed effects for administrative units and regional seasonality, with standard errors clustered at the administrative level.

**Figure S9:**
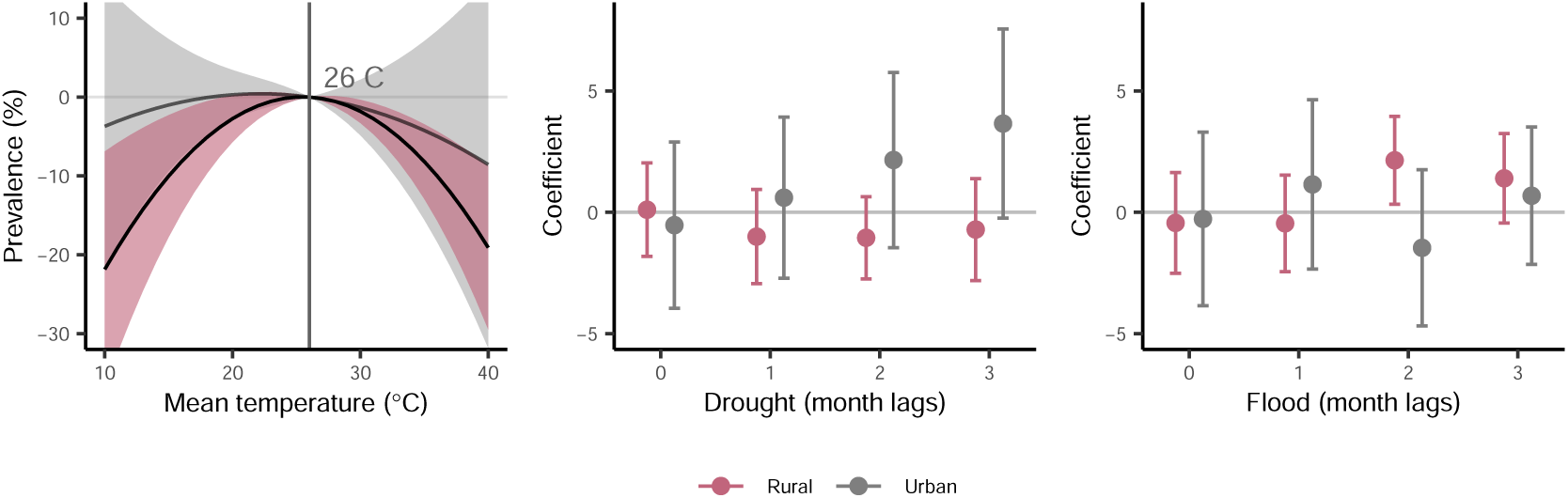
Heterogeneous effects of temperature and extreme precipitation on malaria prevalence by urban and rural status. All panels show the estimated relationship between malaria prevalence for children aged 2-10 estimated separately for rural (pink) and urban (gray) areas. (Left) The relationship between prevalence and mean monthly temperature with shading indicating 95% confidence intervals. (Middle, Right) Estimated coefficients and 95% confidence intervals for the effects of drought (middle) and flood (right) events across contemporaneous and lagged months. Positive coefficients indicate higher malaria prevalence associated with extreme climate events. Model estimates include fixed effects for administrative units and regional seasonality, with standard errors clustered at the administrative level. Urban–rural classifications are based on the Global Human Settlement Layer Urban Centre Database. ^108^

**Figure S10:**
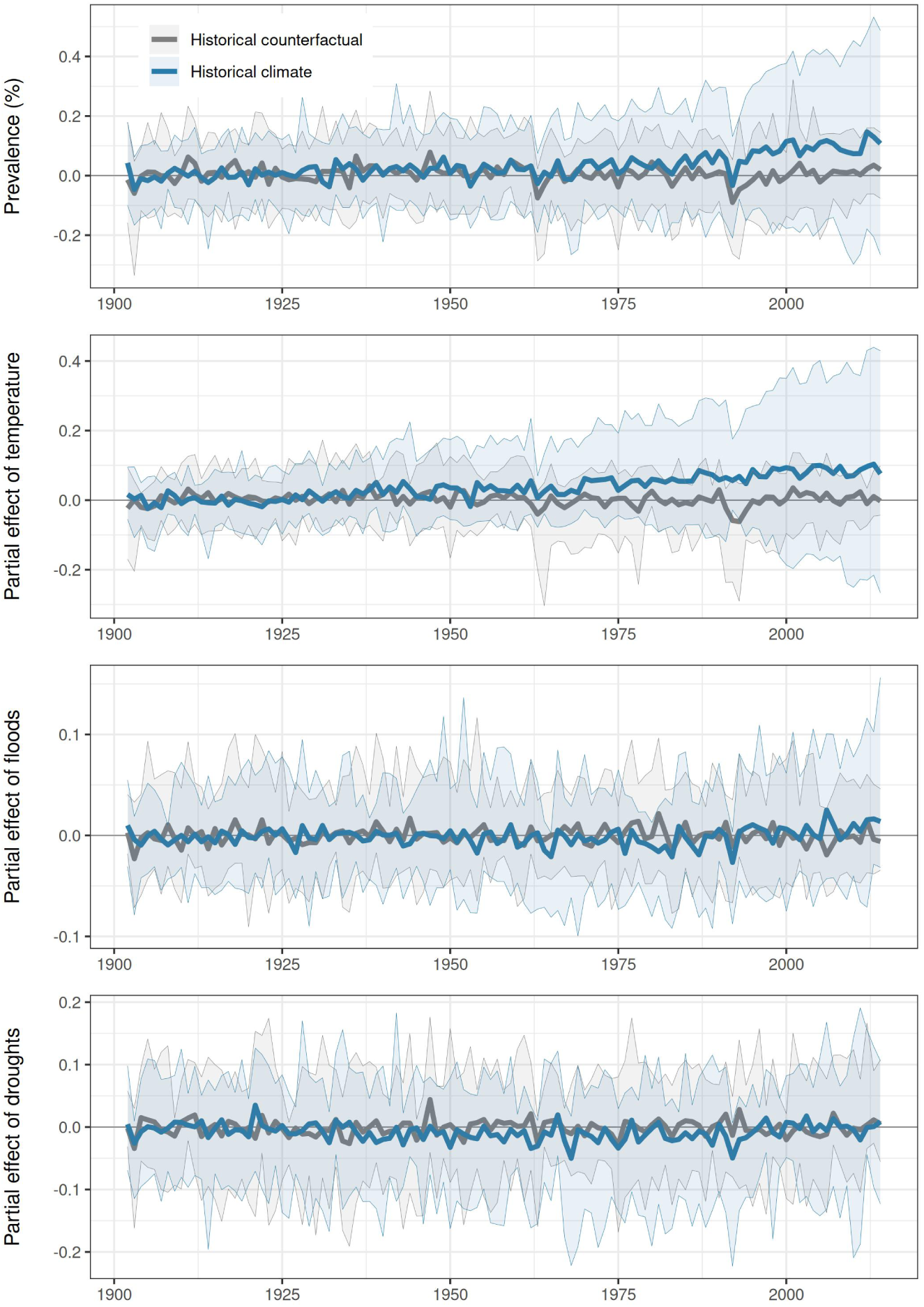
Historical impacts of climate change decomposed by variable. Partial predictions of changes in malaria attributable to anthropogenic climate change are made based on all climate variables (top row), temperature (second row), flood shocks (third row), and drought shocks (fourth row). As in Figure 2, predictions based on true historical climate (blue) are compared to counterfactual predictions without anthropogenic warming (grey), relative to a 1901 to 1930 baseline. Thick lines are the median estimate across all 10,000 simulations; shading indicates the 5*^th^* and 95*^th^* percentiles. Plots begin in 1902 with the first full year of predictions (due to lag effects).

**Figure S11:**
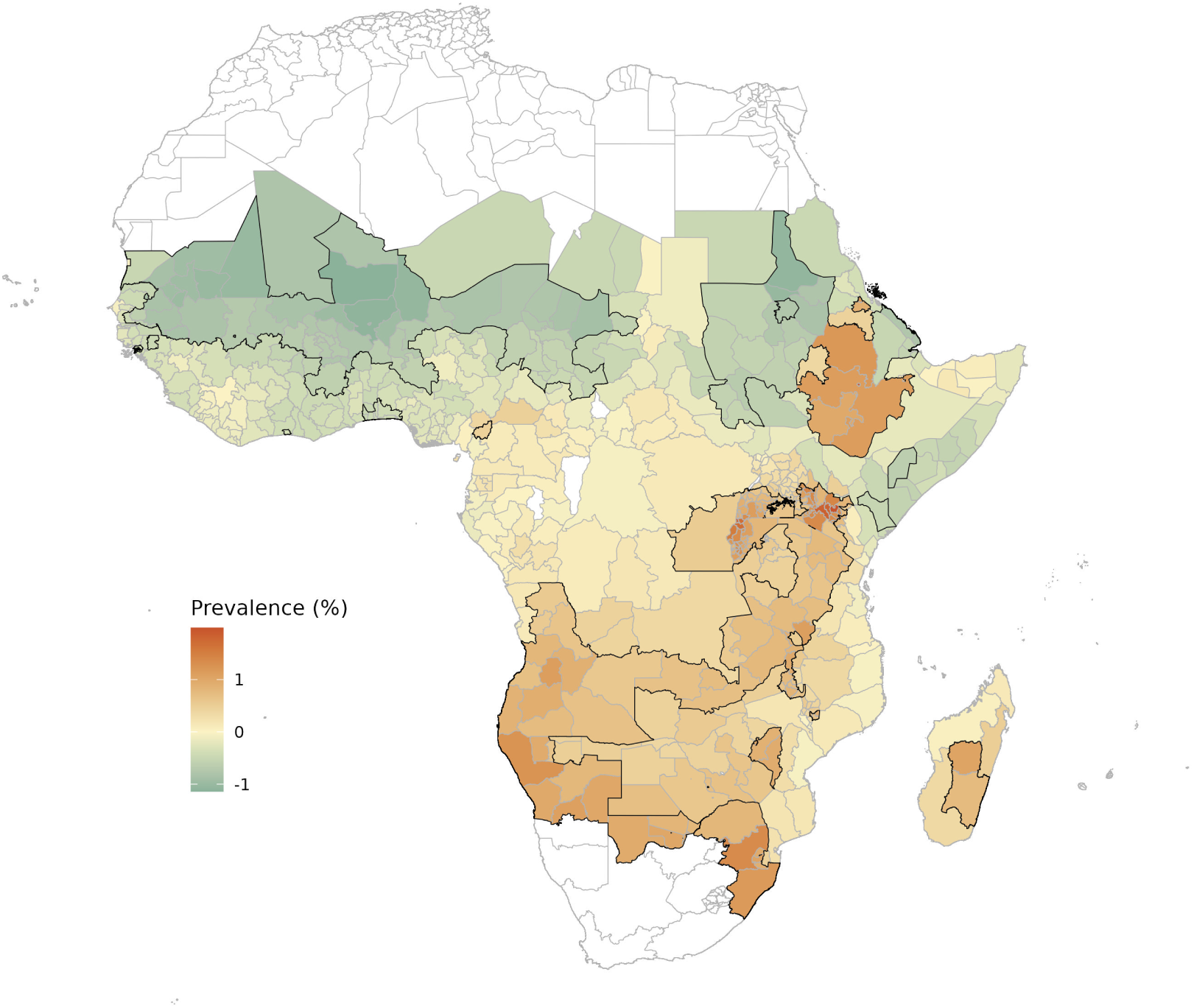
Historical changes in malaria prevalence attributable to anthropogenic climate change from 1901 to 2014. Map shows the estimated change in prevalence attributable to anthropogenic climate change in each administrative unit, based on the difference between the historical climate in 2010-2014 and a counterfactual scenario for the same period simulated without anthropogenic warming. Polygons with a black solid outline indicate areas with changes that were statistically significant (*α* = 0.05) based on the sign of 10,000 boot-strapped simulations. Mean estimates shown here provide the same information as in Figure 3, but on a single color scale (*i.e.*, no uncertainty visualization).

**Figure S12:**
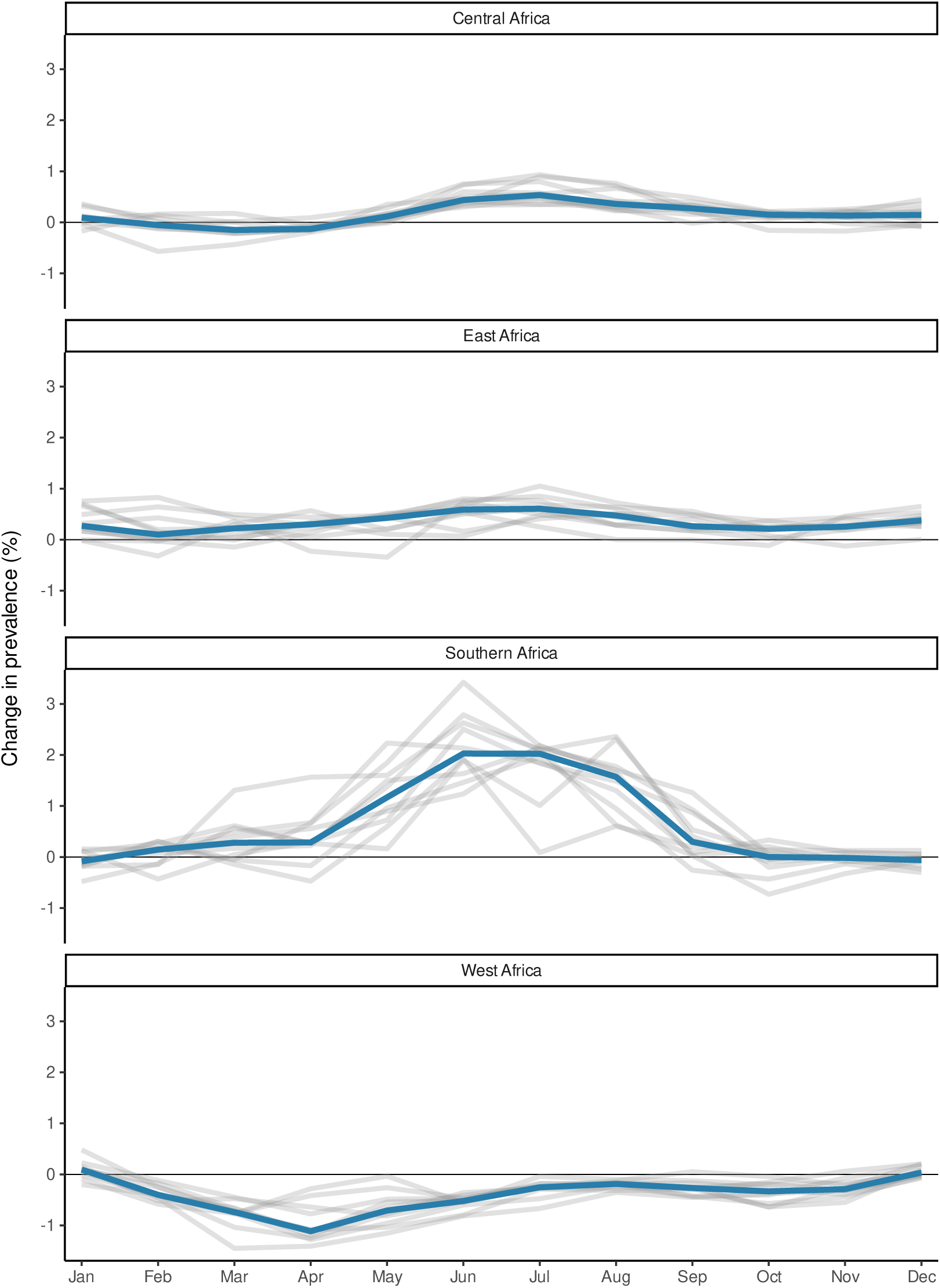
Seasonality in the impacts of anthropogenic climate change on malaria prevalence. Line plots show, for each Global Burden of Disease region, the difference between predicted monthly prevalence under the true historical climate and a counterfactual climate without anthropogenic warming. Positive values indicate that historical climate change elevated malaria prevalence during the indicated month. Impacts are shown as averages by month over the last 5 years (2010-2014) of the historical sample. Each grey line represents one climate model projection, with the blue line showing the median across all models. Associated tabular summaries are shown in Table S3.

**Figure S13:**
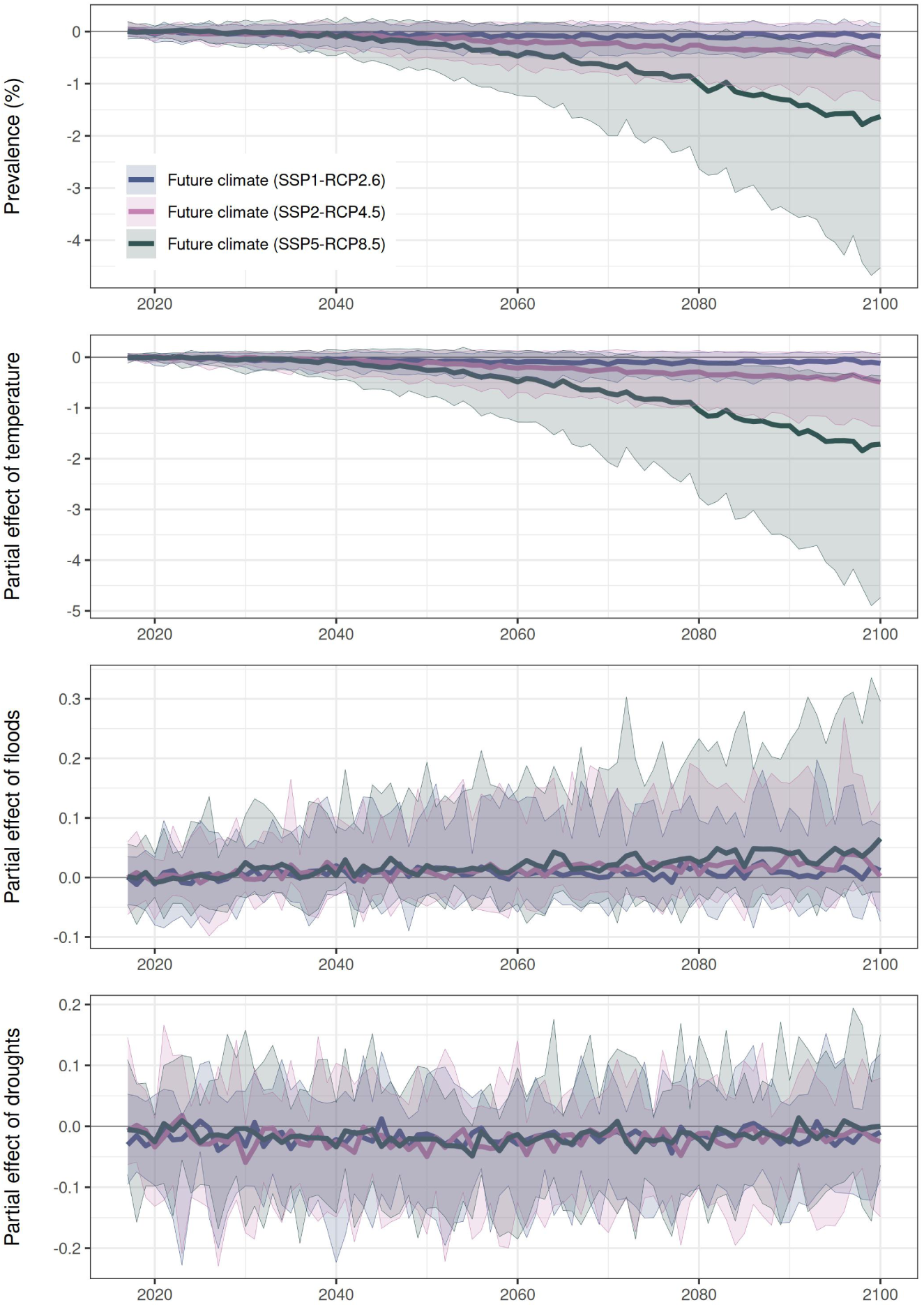
Future impacts of climate change decomposed by variable. Partial predictions of changes in malaria attributable to future climate change are made based on all climate variables (top row) temperature (second row), flood shocks (third row), and drought shocks (fourth row). Projections are shown relative to the mean prevalence from 2015-2020, and as in Figure 2, line color indicates emissions scenario (blue: SSP1-RCP2.6; pink: SSP2-RCP4.5; green: SSP5-RCP8.5). Thick lines are the median estimate across all 10,000 simulations; shading indicates the 5*^th^* and 95*^th^* percentiles. Plots begin in 2016 with the first full year of predictions (due to lag effects).

**Figure S14:**
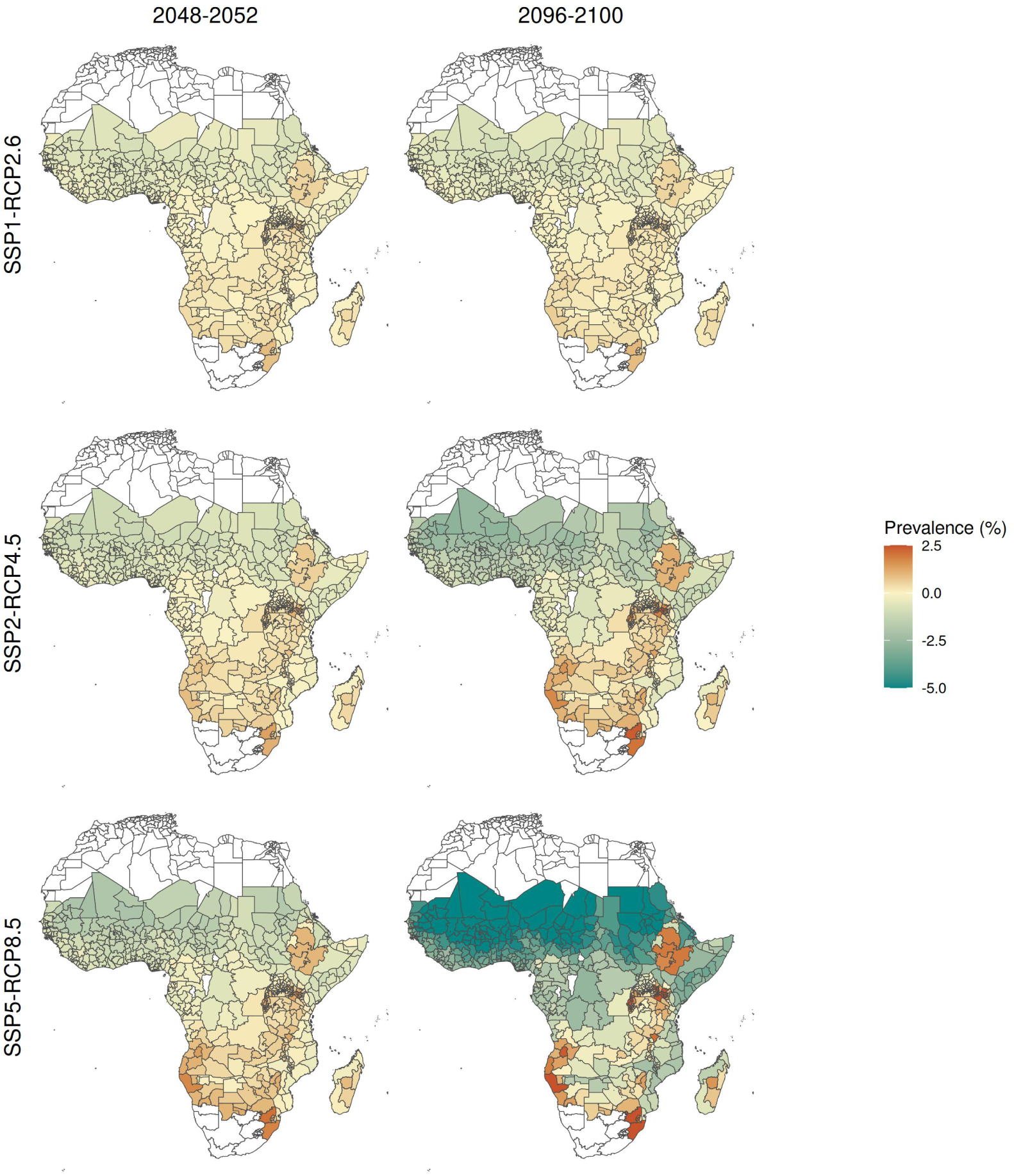
Projected future changes in malaria prevalence driven by climate change from 2015 to 2100. Maps show the estimated change in prevalence due to anthropogenic climate change (in percentage points) in low-emissions (SSP1-RCP2.6; row 1), moderate-emissions (SSP2-RCP4.5; row 2), and high-emissions (SSP5-RCP8.5; row 3) scenarios, projected to mid-century (2048-2052; column 1) or the end of the century (2096-2100; column 2). Projections are reported as differences relative to a present-day baseline (2015-2019). Mean estimates in row 2 column 2 provide the same information as in Figure 4, but on a single color scale (*i.e.*, no uncertainty visualization). The color bar is winsorized for display purposes only, the full range across all regions is -8.1 *p.p.* to 5.5 *p.p.*.

**Figure S15:**
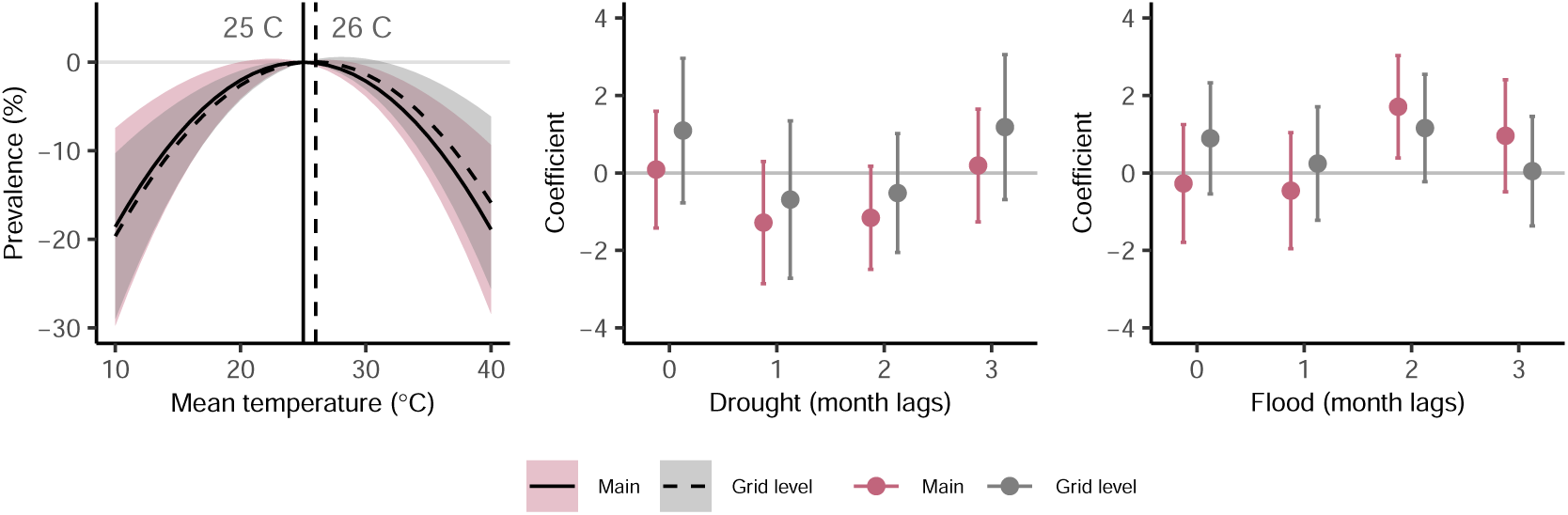
Empirical estimates of climate-prevalence relationships at administrative versus grid cell spatial resolution. Panels show the estimated relationship between malaria prevalence for children aged 2-10 and monthly average temperature (left), drought (middle), and flood (right), estimated separately for our main model specification, aggregated to the first level of administrative division, and for a high-resolution model aggregated to the CRU climate data grid (resolution 0.5*^◦^*). (Left) The relationship between prevalence and mean monthly temperature for the main ADM1 model (black solid line and pink shading) and high resolution grid-level model (dashed line and grey shading), with shading indicating 95% confidence intervals. (Middle, right) Estimated coefficients and 95% confidence intervals for the effects of drought (middle) and flood (right) events across contemporaneous and lagged months for both model specifications. Positive coefficients indicate higher malaria prevalence associated with extreme climate events. Models are estimated following Equation 4 (main) and Equation 5 (grid level) and all standard errors are clustered at the administrative level (ADM1).

**Figure S16:**
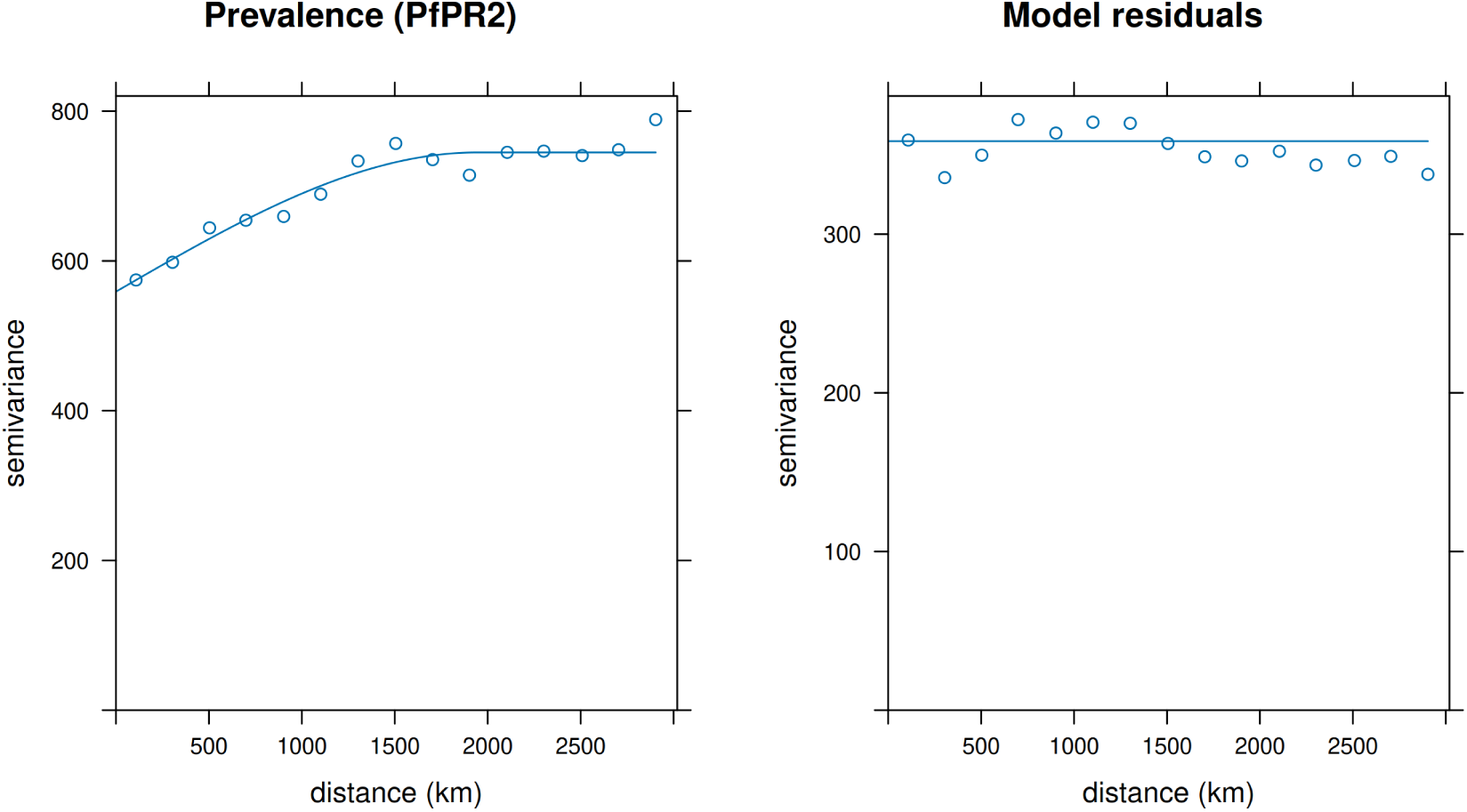
Empirical variogram of malaria prevalence and regression model residuals. Each panel shows estimated semi-variances (*y*-axis) against distance between spatial units (*x*-axis). Semi-variances are defined for each distance *h* as half the variance of the difference between values located within *h* units of each other ^106^. When spatial correlation is present, semi-variances rise with distance, as variance grows with the distance between observations. (Left) The variogram for malaria prevalence *Pf* PR_2_*_−_*_10_. (Right) The variogram for regression model residuals, as estimated following Equation 4. Hollow circles indicate binned scatters while solid lines indicate a spherical variogram model fit. All estimates derived using the gstat package in R.

**Figure S17:**
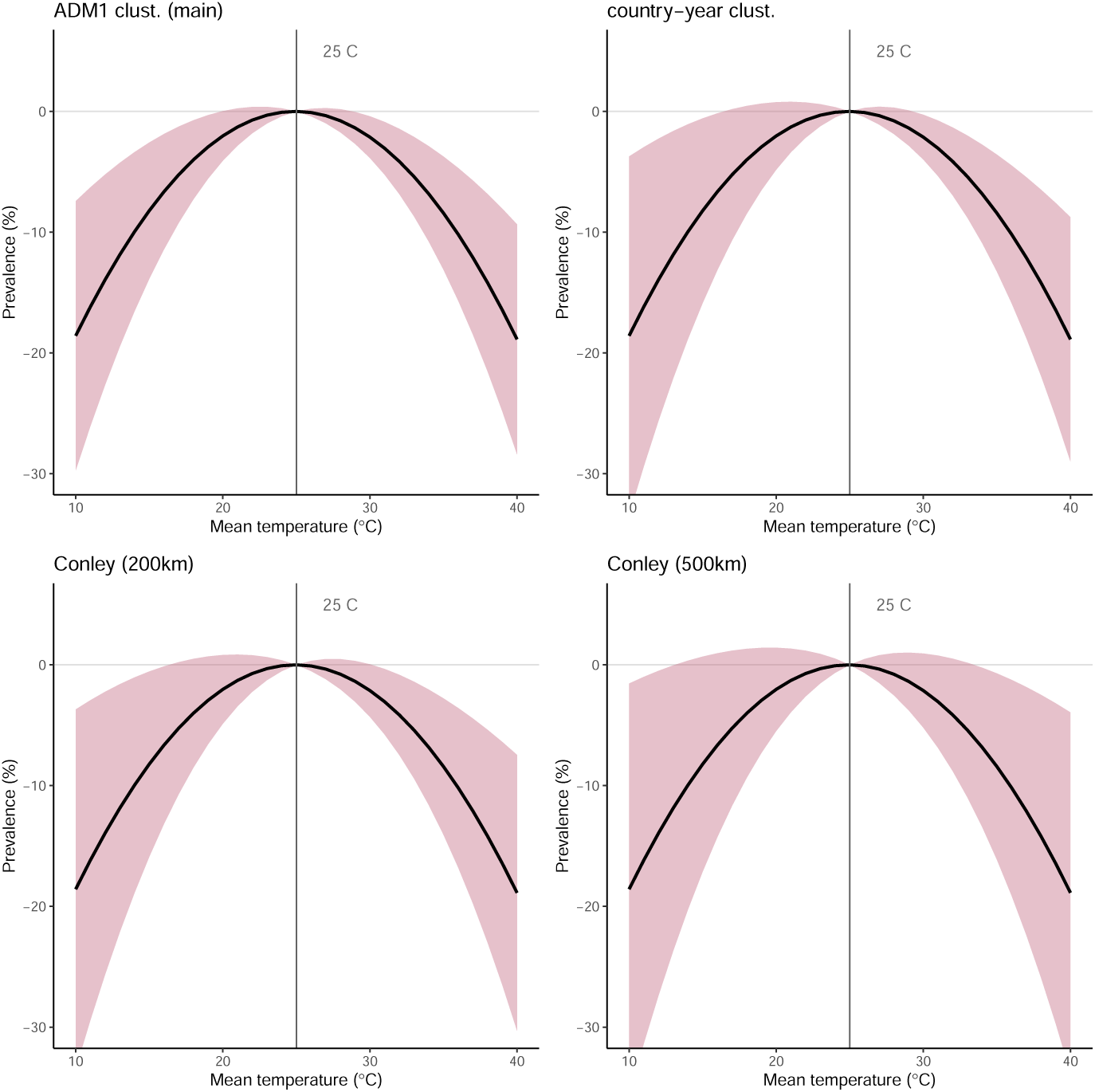
Alternative estimates of model uncertainty in the *Pf* **PR**_2_*_−_*_10_-temperature relationship. All panels show the estimated relationship between malaria prevalence for children aged 2-10 and monthly average temperature as estimated following Equation 4. All temperature responses are plotted relative to 25*^◦^*C, the temperature at which prevalence is maximized. From top-left to bottom-right, standard errors are estimated using: ADM1-level clustering (main specification), country-by-year level clustering, Conley standard errors with a cutoff of 200 km, and Conley standard errors with a cutoff of 500 km. Tabular results, including associated estimates of precipitation extremes, are shown in Table S6.

**Figure S18:**
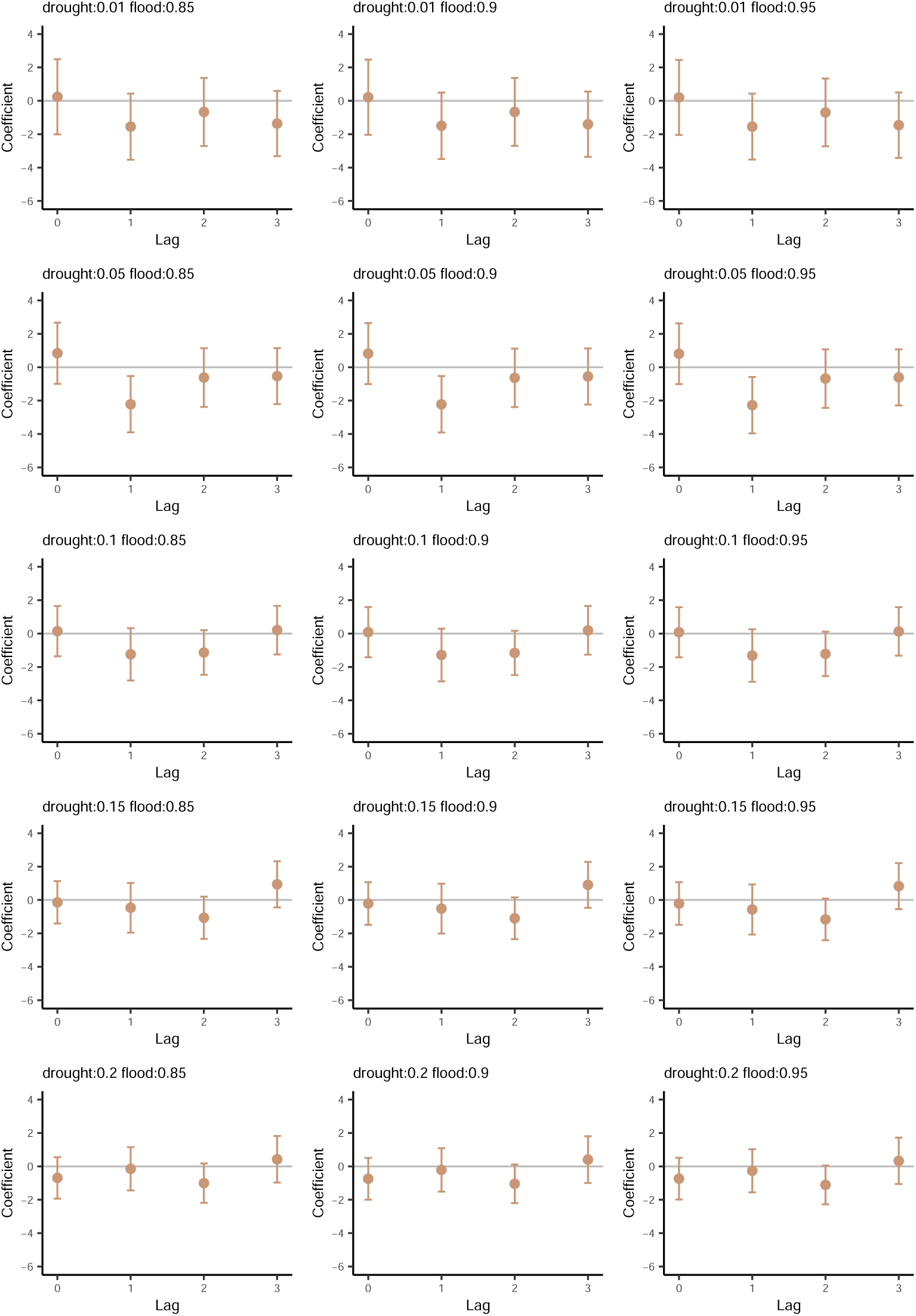
Sensitivity of *Pf* PR_2_*_−_*_10_-drought relationship to alternative drought and flood definitions. All panels show the estimated relationship between malaria prevalence for children aged 2-10 and contemporaneous and lagged drought events. Point estimates are given by solid circles, while vertical bars indicate 95% confidence intervals. Cutoff values for drought and flood definitions are given in the titles of each panel. For example, the first panel in the upper left defines drought as monthly total precipitation that falls below 1% of the long-run location- and month-specific mean, and defines flood as monthly total precipitation that falls above 85% of the long-run location- and month-specific mean. In all specifications, three months of lagged precipitation extremes are included, as well as all other controls shown in Equation 4. The main specification used throughout the paper uses a drought cutoff of 10% and a flood cutoff of 90%. All standard errors are clustered at the first administrative unit (e.g., province) level.

### Supplementary Tables

**Table S1:** Sensitivity of estimated effects of temperature and rainfall on *Pf* PR_2–10_ to controlling for malaria diagnostic method. All columns show regression results for a dependent variable of malaria prevalence among children aged 2–10 (*Pf* PR_2–10_). Column (1) reproduces the preferred specification from Table S2, column (5) with a single observation that uses loop-mediated isothermal amplification (LAMP) dropped to make for a direct comparison with columns (2) and (3). Column (2) adds a full set of indicators for the dominant diagnostic method observed in each ADM1–year–month: Microscopy–PCR confirmed, PCR, RDT, RDT–PCR confirmed, RDT–Slide confirmed, and Slide confirmed (Microscopy is the omitted category; a single LAMP observation is dropped). Column (3) replaces these with a simplified method classification that retains only PCR and RDT indicators, again leaving Microscopy as the reference and leaving out the LAMP observation. Standard errors are clustered at the first administrative unit (ADM1) level.

|  | (1) | (2) | (3) |
| --- | --- | --- | --- |
| Avg temp. (°C) | 4.15***<br>(1.08) | 3.97***<br>(1.09) | 4.08***<br>(1.09) |
| Avg temp.<sup>2</sup> (°C) | -0.08***<br>(0.02) | -0.08***<br>(0.02) | -0.08***<br>(0.02) |
| Flood | -0.28<br>(0.78) | -0.06<br>(0.78) | -0.22<br>(0.78) |
| Flood (lag 1) | -0.45<br>(0.76) | -0.45<br>(0.77) | -0.47<br>(0.76) |
| Flood (lag 2) | 1.70**<br>(0.68) | 1.81***<br>(0.67) | 1.83***<br>(0.67) |
| Flood (lag 3) | 0.96<br>(0.74) | 1.05<br>(0.74) | 0.97<br>(0.74) |
| Drought | 0.09<br>(0.77) | 0.18<br>(0.76) | 0.23<br>(0.77) |
| Drought (lag 1) | -1.28<br>(0.80) | -1.38*<br>(0.81) | -1.28<br>(0.80) |
| Drought (lag 2) | -1.14*<br>(0.68) | -1.18*<br>(0.68) | -1.15*<br>(0.68) |
| Drought (lag 3) | 0.19<br>(0.74) | 0.09<br>(0.75) | 0.15<br>(0.75) |
| Intvn. 1955-69 | -4.82***<br>(1.27) | -4.98***<br>(1.28) | -4.76***<br>(1.27) |
| Intvn. 2000-15 | -3.37**<br>(1.61) | -3.19**<br>(1.60) | -3.60**<br>(1.60) |
| Diag. Method Microscopy - PCR Confirmed |  | 6.13<br>(4.75) |  |
| Diag. Method PCR |  | 27.86***<br>(3.86) |  |
| Diag. Method RDT |  | 2.14<br>(1.35) |  |
| Diag. Method RDT - PCR Confirmed |  | 10.29<br>(11.04) |  |
| Diag. Method RDT - Slide Confirmed |  | -9.02***<br>(1.34) |  |
| Diag. Method (simplified) PCR |  |  | 27.62***<br>(3.85) |
| Diag. Method (simplified) RDT |  |  | -0.60<br>(1.12) |
| Observations | 9,874 | 9,874 | 9,874 |
| R<sup>2</sup> | 0.53 | 0.53 | 0.53 |
| Adjusted R<sup>2</sup> | 0.49 | 0.50 | 0.50 |

**Table S2:** Sensitivity of estimated effects of temperature and rainfall on *Pf* PR_2_*_−_*_10_ to alternative spatiotemporal controls. All columns show regression results for a dependent variable of malaria prevalence for children aged 2-10 (*Pf* PR_2_*_−_*_10_). All include fixed effects (i.e., dummy variables) at the scale of the first administrative unit (i.e., ADM1). Other model controls by column are: (1) country, year, and month fixed effects; (2) country-specific quadratic time trends and month fixed effects; (3) country-specific quadratic time trends and country-by-month fixed effects; (4) country-specific quadratic time trends and intervention period and month fixed effects; (5) country-specific quadratic time trends and intervention period and region-by-month fixed effects; (6) country-specific quadratic time trends and intervention period and country-by-month fixed effects; (7) region-by-year and region-by-month fixed effects; (8) region-by-year and country-by-month fixed effects; (9) and region-by-year and region-by-month fixed effects and country-specific linear time trends. The preferred specification used throughout the main text is column (5). In columns (5) and (7)-(9), “region” refers to the Global Burden of Disease regional definitions of western, southern, central, and eastern Africa. All standard errors are clustered at the first administrative unit (e.g., province) level.

| *Dependent variable:* |  |  |  |  |  |  |  |  |  |
| --- | --- | --- | --- | --- | --- | --- | --- | --- | --- |
|  | cnty + yr<br>+ mo FEs.<br>(1) | cnty trd,<br>mo FEs.<br>(2) | cnty trd,<br>cnty-mo FEs.<br>(3) | cnty trd,<br>int + mo FEs.<br>(4) | cnty trd,<br>int + rgn-mo FEs.<br>(5) | cnty trd,<br>int +<br>cnty-mo FEs.<br>(6) | rgn-yr +<br>rgn-mo FEs.<br>(7) | rgn-yr +<br>cnty-mo FEs.<br>(8) | rgn-yr + rgn-mo FEs,<br>cnty trd<br>(9) |
| *PfPR<sub>2-10</sub>* |  |  |  |  |  |  |  |  |  |
| Avg temp. (°C) | 2.42***<br>(0.83) | 3.08***<br>(0.88) | 3.80***<br>(1.39) | 3.23***<br>(0.87) | 4.15***<br>(1.08) | 3.97***<br>(1.39) | 3.13***<br>(1.00) | 2.11<br>(1.34) | 3.06***<br>(0.99) |
| Avg temp.<sup>2</sup> (°C) | -0.06***<br>(0.02) | -0.07***<br>(0.02) | -0.07***<br>(0.03) | -0.07***<br>(0.02) | -0.08***<br>(0.02) | -0.08***<br>(0.03) | -0.07***<br>(0.02) | -0.05*<br>(0.03) | -0.06***<br>(0.02) |
| Flood | -0.76<br>(0.75) | -0.37<br>(0.74) | -0.73<br>(0.82) | -0.27<br>(0.74) | -0.27<br>(0.78) | -0.65<br>(0.82) | -0.88<br>(0.76) | -1.15<br>(0.80) | -0.82<br>(0.74) |
| Flood (lag 1) | -0.56<br>(0.73) | -0.40<br>(0.76) | -0.50<br>(0.85) | -0.46<br>(0.75) | -0.46<br>(0.76) | -0.55<br>(0.83) | -0.77<br>(0.73) | -1.09<br>(0.80) | -0.57<br>(0.72) |
| Flood (lag 2) | 1.68***<br>(0.65) | 1.52**<br>(0.65) | 2.03***<br>(0.72) | 1.69***<br>(0.65) | 1.71**<br>(0.68) | 2.19***<br>(0.72) | 2.13***<br>(0.68) | 1.94***<br>(0.72) | 1.79***<br>(0.67) |
| Flood (lag 3) | 0.58<br>(0.69) | 0.90<br>(0.71) | 1.89**<br>(0.80) | 0.87<br>(0.71) | 0.96<br>(0.74) | 1.91**<br>(0.81) | 0.08<br>(0.70) | 0.74<br>(0.77) | 0.38<br>(0.68) |
| Drought | 0.56<br>(0.81) | 0.87<br>(0.74) | -0.48<br>(0.82) | 0.84<br>(0.74) | 0.09<br>(0.77) | -0.57<br>(0.83) | -0.35<br>(0.81) | -0.69<br>(0.86) | -0.13<br>(0.74) |
| Drought (lag 1) | -1.01<br>(0.75) | -1.02<br>(0.77) | -1.39<br>(0.86) | -0.92<br>(0.78) | -1.28<br>(0.80) | -1.34<br>(0.86) | -1.62**<br>(0.76) | -1.28<br>(0.85) | -1.33*<br>(0.77) |
| Drought (lag 2) | -0.97<br>(0.68) | -1.08*<br>(0.65) | -0.88<br>(0.76) | -1.04<br>(0.65) | -1.16*<br>(0.68) | -0.95<br>(0.76) | -1.18*<br>(0.69) | -0.85<br>(0.77) | -1.24*<br>(0.67) |
| Drought (lag 3) | 0.06<br>(0.71) | 0.16<br>(0.73) | 0.36<br>(0.81) | 0.06<br>(0.73) | 0.19<br>(0.74) | 0.15<br>(0.80) | -0.06<br>(0.77) | 0.11<br>(0.83) | -0.16<br>(0.75) |
| Intvn. 1955-69 |  |  |  |  | -4.80***<br>(1.27) | -5.01***<br>(1.35) |  |  |  |
| Intvn. 2000-15 |  |  |  |  | -3.35**<br>(1.61) | -3.88**<br>(1.64) |  |  |  |
| Observations | 9,875 | 9,875 | 9,875 | 9,875 | 9,875 | 9,875 | 9,875 | 9,875 | 9,875 |
| R<sup>2</sup> | 0.52 | 0.52 | 0.56 | 0.53 | 0.53 | 0.57 | 0.56 | 0.60 | 0.59 |
| Adjusted R<sup>2</sup> | 0.48 | 0.49 | 0.51 | 0.49 | 0.49 | 0.51 | 0.52 | 0.53 | 0.54 |

**Table S3:** Seasonal peaks in the impacts of historical anthropogenic climate change on malaria prevalence. Table summarizes key statistics from line plots in Figure S12. For each Global Burden of Disease region, the month of peak impact indicates the month with the largest absolute difference between predicted prevalence with (historical true climate) and without (counterfactual climate) anthropogenic climate change. Impact size represents the increase or decrease in predicted prevalence due to climate change during the month of peak impact, averaged over the 2010-2014 time period. 95% confidence intervals are shown in parentheses. For comparison, average annual impacts and their confidence intervals are shown in the last column (these values correspond to those shown at the end of the annual time series for each region in Figure 3D).

| Region | Month of<br>Peak Impact | Impact Size<br>(% points) | Average Annual<br>Impact (% points) |
| --- | --- | --- | --- |
| Central Africa | Jul | 0.58 (-0.13, 1.52) | 0.18 (-0.19, 0.61) |
| East Africa | Jul | 0.65 (-0.26, 1.76) | 0.33 (-0.14, 0.87) |
| Southern Africa | Jun | 2.26 (0.35, 4.71) | 0.63 (-0.04, 1.40) |
| West Africa | Apr | -0.92 (-2.35, 0.13) | -0.38 (-0.90, 0.02) |

**Table S4:** Projected future impacts of climate change on *Pf* PR_2_*_−_*_10_. Estimates and confidence intervals are all given as percentage point changes from a 2015-2020 baseline, estimated across 10,000 simulations. P_+_ indicates the proportion of simulations showing an increase in malaria prevalence relative to the baseline period.

| Region | Scenario | 2048-2052 |  |  | 2096-2100 |  |  |
| --- | --- | --- | --- | --- | --- | --- | --- |
|  |  | Estimate | 95% CI | P<sub>+</sub> | Estimate | 95% CI | P<sub>+</sub> |
| Sub-Saharan Africa<br>(continent-wide) | SSP1-RCP2.6 | -0.086 | (-0.411, 0.169) | 0.254 | -0.094 | (-0.497, 0.160) | 0.311 |
|  | SSP2-RCP4.5 | -0.148 | (-0.570, 0.204) | 0.223 | -0.433 | (-1.371, 0.249) | 0.122 |
|  | SSP5-RCP8.5 | -0.244 | (-0.803, 0.261) | 0.186 | -1.893 | (-4.846, -0.065) | 0.021 |
| East Africa | SSP1-RCP2.6 | 0.112 | (-0.179, 0.466) | 0.774 | 0.098 | (-0.248, 0.449) | 0.806 |
|  | SSP2-RCP4.5 | 0.131 | (-0.268, 0.543) | 0.750 | 0.131 | (-0.577, 0.846) | 0.655 |
|  | SSP5-RCP8.5 | 0.157 | (-0.371, 0.688) | 0.729 | -0.603 | (-2.715, 0.951) | 0.266 |
| Central Africa | SSP1-RCP2.6 | -0.035 | (-0.354, 0.205) | 0.427 | -0.060 | (-0.430, 0.242) | 0.390 |
|  | SSP2-RCP4.5 | -0.021 | (-0.471, 0.385) | 0.482 | -0.231 | (-1.192, 0.514) | 0.302 |
|  | SSP5-RCP8.5 | -0.109 | (-0.690, 0.444) | 0.357 | -1.433 | (-4.361, 0.371) | 0.078 |
| Southern Africa | SSP1-RCP2.6 | 0.321 | (-0.094, 0.836) | 0.939 | 0.339 | (-0.062, 0.925) | 0.945 |
|  | SSP2-RCP4.5 | 0.507 | (-0.055, 1.104) | 0.962 | 0.844 | (-0.185, 1.940) | 0.944 |
|  | SSP5-RCP8.5 | 0.846 | (-0.079, 1.810) | 0.962 | 0.492 | (-2.313, 2.547) | 0.703 |
| West Africa | SSP1-RCP2.6 | -0.448 | (-0.952, -0.135) | 0.002 | -0.449 | (-1.087, 0.065) | 0.096 |
|  | SSP2-RCP4.5 | -0.694 | (-1.290, -0.232) | 0.001 | -1.510 | (-3.045, -0.481) | 0.001 |
|  | SSP5-RCP8.5 | -1.041 | (-1.860, -0.358) | 0.001 | -4.246 | (-8.780, -1.519) | 0.000 |

**Table S5:** Summary statistics of temporal and spatial correlations of model residuals. Values show mean correlation (*ρ*), interquartile range (Q25, Q75), and sample size (*N*) for different groupings temporally (Kind = “temporal”) or spatially (Kind = “spatial”), following ref. ^105^. For temporal groupings, correlations between month-year observations within the same ADM1 unit are estimated. For spatial groupings, correlations between month-year sequences of residuals from different ADM1 units are estimated. Here, “region” indicates one of four Global Burden of Disease regions. Means, first quartiles, and third quartiles of the distribution of correlations from each kind of grouping are shown. *N* indicates the number of pairs for which correlations could be estimated; pairs with less than 10 matching observations were dropped.

| Kind | Group | $\bar{\rho}$ | Q25 | Q75 | $N$ |
| --- | --- | --- | --- | --- | --- |
| temporal | all | 0.18 | -0.09 | 0.48 | 511 |
| temporal | 1 month lag | 0.40 | 0.20 | 0.65 | 60 |
| temporal | 2 month lag | 0.17 | -0.06 | 0.43 | 33 |
| temporal | 3 month lag | 0.22 | -0.05 | 0.43 | 16 |
| temporal | same month, different year | 0.22 | -0.04 | 0.53 | 41 |
| spatial | all | 0.24 | -0.01 | 0.54 | 635 |
| spatial | same country | 0.28 | 0.04 | 0.56 | 537 |
| spatial | different country | 0.04 | -0.18 | 0.28 | 98 |
| spatial | same region | 0.04 | -0.17 | 0.19 | 51 |
| spatial | different region | 0.04 | -0.19 | 0.30 | 47 |
| spatial | < 500km | 0.30 | 0.04 | 0.60 | 395 |
| spatial | > 500km | 0.15 | -0.10 | 0.39 | 240 |
| spatial | > 1000km | 0.07 | -0.17 | 0.33 | 103 |

**Table S6:**
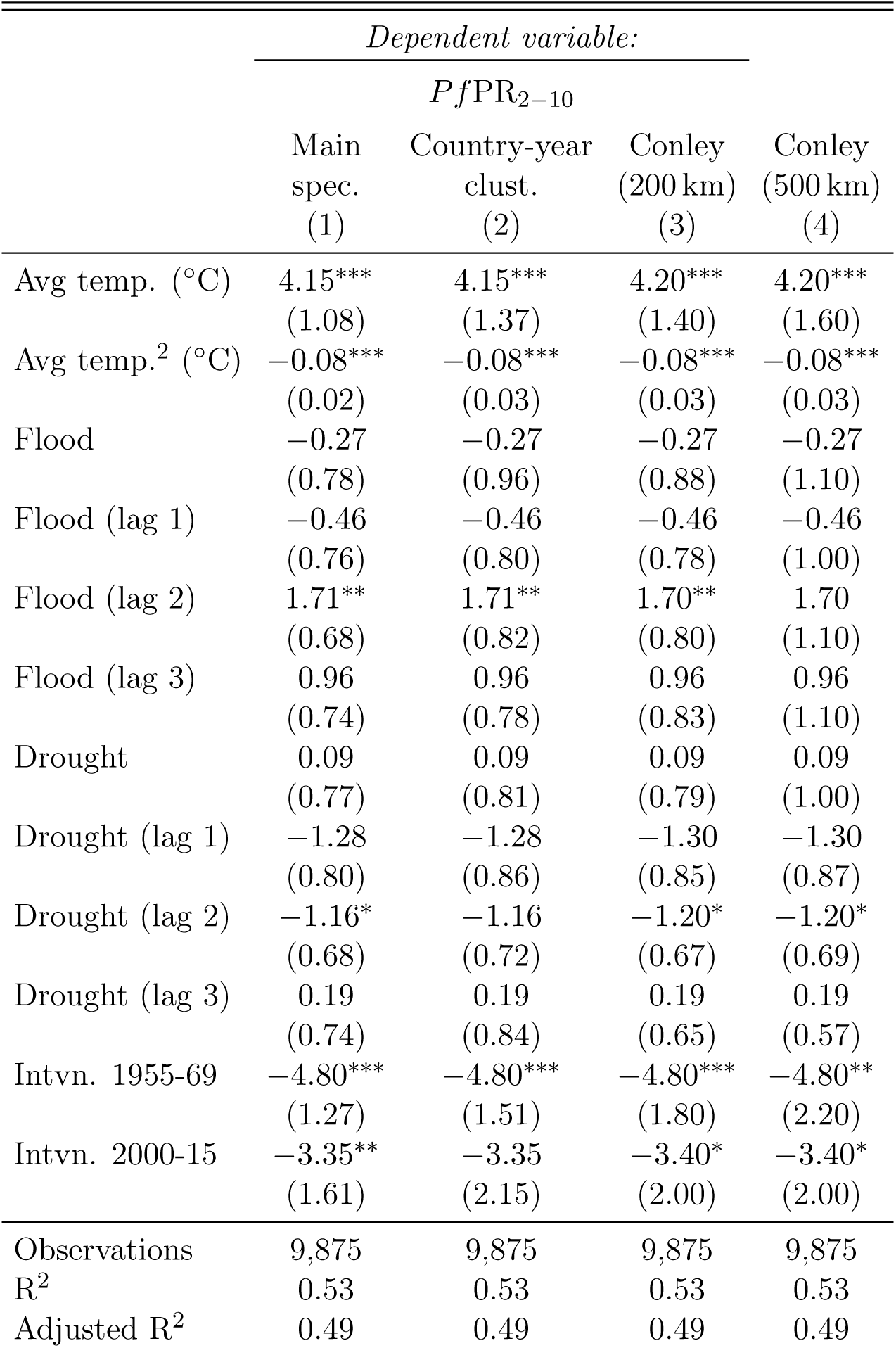
Alternative estimates of model uncertainty in *Pf* PR_2_*_−_*_10_-weather relationships. All columns show regression results for a dependent variable of malaria prevalence for children aged 2-10 (*Pf* PR_2_*_−_*_10_) and all are estimated following Equation 4. In each column, standard errors are estimated under different assumptions of spatial and/or temporal correlation in model residuals. Specifications are: ADM1-level clustering (column 1, main specification); country-by-year level clustering (column 2); Conley standard errors with a cutoff of 200 km (column 3); and Conley standard errors with a cutoff of 500 km (column 4). Figure S17 plots the resulting temperature response functions and associated uncertainty.

